# Progressive accumulation of hyperinflammatory NKG2D^low^ NK cells in early childhood severe atopic dermatitis

**DOI:** 10.1101/2023.06.02.23290884

**Authors:** David E. Ochayon, Stanley B. DeVore, Wan-Chi Chang, Durga Krishnamurthy, Harsha Seelamneni, Brittany Grashel, Daniel Spagna, Sandra Andorf, Lisa J. Martin, Jocelyn M. Biagini, Stephen N. Waggoner, Gurjit K. Khurana Hershey

## Abstract

Atopic dermatitis (AD) is a chronic inflammatory skin disease that often precedes the development of food allergy, asthma, and allergic rhinitis. The prevailing paradigm holds that a reduced frequency and function of natural killer (NK) cell contributes to AD pathogenesis, yet the underlying mechanisms and contributions of NK cells to allergic comorbidities remain ill-defined. Herein, analysis of circulating NK cells in a longitudinal early life cohort of children with AD revealed a progressive accumulation of NK cells with low expression of the activating receptor NKG2D, which was linked to more severe AD and sensitivity to allergens. This was most notable in children co-sensitized to food and aero allergens, a risk factor for development of asthma. Individual-level longitudinal analysis in a subset of children revealed co-incident reduction of NKG2D on NK cells with acquired or persistent sensitization, and this was associated with impaired skin barrier function assessed by transepidermal water loss. Low expression of NKG2D on NK cells was paradoxically associated with depressed cytolytic function but exaggerated release of the proinflammatory cytokine TNF−α. These observations provide important insights into a potential mechanism underlying the development of allergic co-morbidity in early life in children with AD which involves altered NK-cell functional responses, and define an endotype of severe AD.

## Introduction

Atopic dermatitis (AD) is an inflammatory skin disease affecting up to 25% of children that often precedes the development of other allergic disorders, including food allergy and allergic rhinitis (*1, 2*). Moreover, nearly half of pediatric patients with AD will develop asthma (*3*). The mechanisms by which AD favors progression to allergies and asthma remain unclear.

Type 2 cytokines (e.g., IL-13) and effector cells (e.g., eosinophils) are generally considered drivers of AD and allergic disease, whereas type 1 immune cells such as natural killer (NK) cells are implicated in a counter regulatory role (*4*). Consistent with this, several studies have contributed a paradigm holding that reduced frequencies and/or diminished cytolytic functionality of circulating NK cells contributes to the pathogenesis of AD (*5-8*). Indeed, IL-15-based therapies that can expand NK cells provided some measure of benefit in mouse models of AD (*5*). NK cells are thought to limit eczema and allergic inflammation via IFN-γ counteraction of type 2 inflammation (*9*) and/or cytolytic regulation of type 2 immune cells (*10-12*). Nevertheless, transcriptomic analyses reveal a progressive enrichment for NK cells in lesional skin of patients with atopic dermatitis (*13-15*). Potential changes in NK-cell function that may contribute to development of allergic disease in children with AD have not been explored.

The Mechanisms of Progression of Atopic Dermatitis to Asthma in Children (MPAACH) cohort (N=651), the first US based prospective early life cohort of children with AD, was designed to elucidate the mechanistic underpinnings of AD and the atopic march (*16-19*). MPAACH is a longitudinal study where children are seen for annual visits that include questionnaires, a physical examination, and extensive biospecimen collections. Herein, we investigated peripheral NK cells in a subset of MPAACH children longitudinally over sequential annual visits to define phenotypic and functional changes that are related to clinical outcomes. Our findings reveal that more severe AD and sensitivity to allergens is associated with selective loss of NKG2D on NK cells but not other immune cells. Correspondingly, there is a progressive accumulation of NKG2D^low^ CD56^dim^ NK cells that exhibit reduced cytolytic function but elevated secretion of proinflammatory cytokines such as TNF-α. Thus, accumulation of hyperinflammatory NKG2D^low^ NK cells defines an endotype of severe AD characterized by increased sensitivity to allergens.

## Results

### Altered repertoire of NK cells in allergen-sensitized children with AD

Peripheral blood samples from any of the first 3 annual visits of a randomly selected subset (n=124, 39-44 subjects per visit) of MPAACH subjects were utilized to assess NK cells. The demographic and clinical characteristics of the included subjects in comparison to the full MPAACH cohort are shown in **Table 1**. Compared to the full cohort, the randomly selected subset was slightly older, less likely to be black, more likely to be co-sensitized to both food and aeroallergens, and more likely to be diagnosed with food allergy. The incidence of comorbid conditions such as wheezing and allergies/allergic rhinitis increased at later clinical visits in this cohort of children (**Table S1**). **Table 1** also shows a comparison of complete blood counts (CBC) in the overall MPAACH cohort and sub-cohort evaluated in this study. The frequencies of different peripheral immune cells were similar in the overall and sub-study cohorts (**Table 1**). Comparisons of cohort demographics and CBCs by visit are shown in **Tables S1 and S2**, respectively.

**Table 1.**
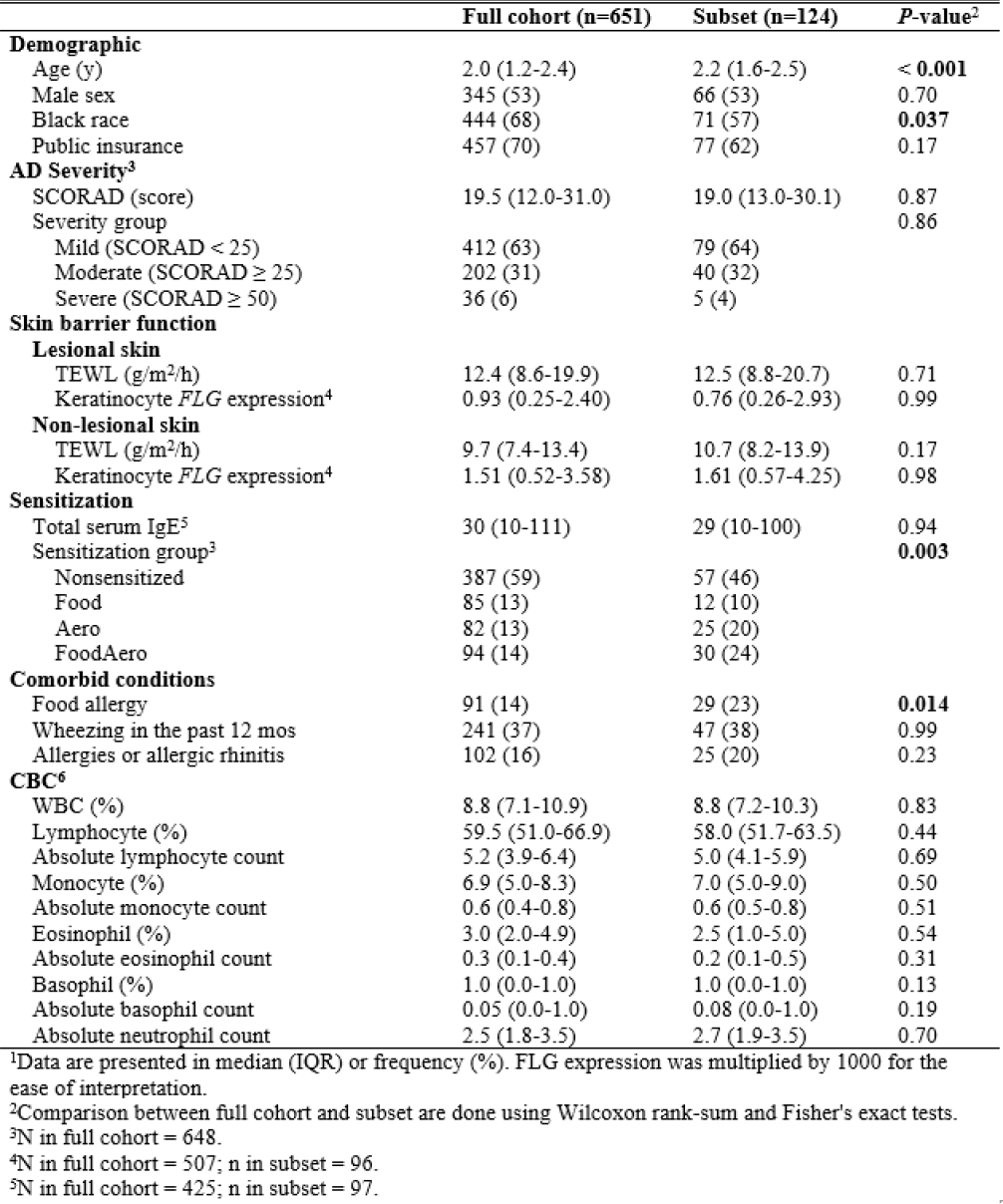
Characteristics of the MPAACH cohort and the Study Subset at Visit I^1^.

Reduced frequencies of NK cells are commonly observed in the blood of patients with AD (*5-8*), but relationships between NK-cell frequencies and comorbidities of AD are undefined. As expected, most NK cells in the peripheral blood of children with AD display a cytolytically mature CD16^+^CD56^dim^ phenotype, with less common populations of more immature CD16^low^CD56^bright^ and CD16^+^CD56^neg^ NK cells (**Fig. S1A+B**). Of note, the frequency of total NK cells (**Fig. 1A**) and the ratio of CD56^dim^ to CD56^bright^ NK cells (**Fig. 1B**) in blood was similar in children with AD across all time points analyzed regardless of allergic sensitivity.

**Figure 1.**
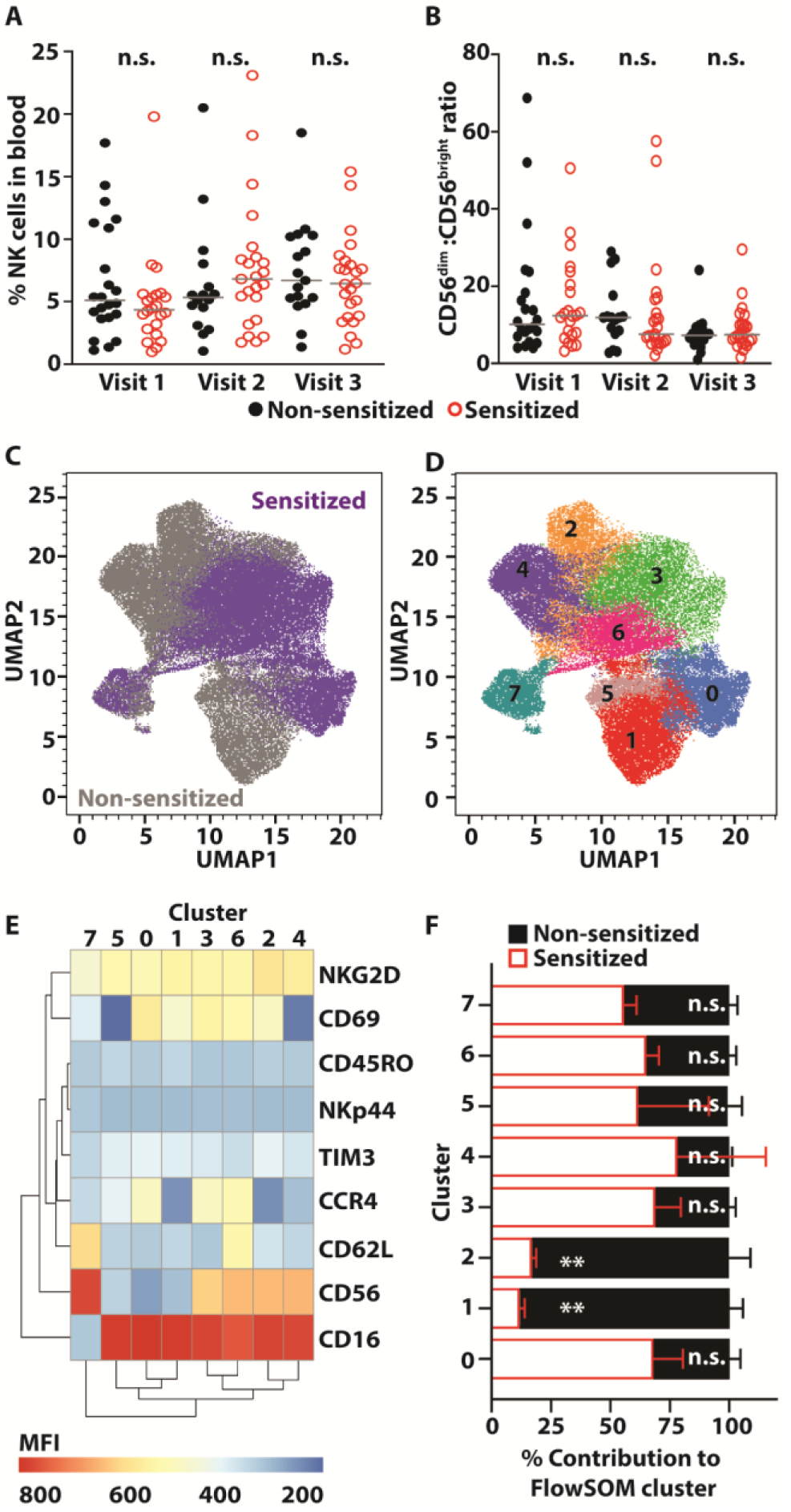
Altered repertoire of NK cells in allergen sensitized children with AD. PBMC from children with AD who lack or exhibit allergic sensitivity (visit 1 (V1)=44, visit 2 (V2)=39, visit 3 (V3), n=41) were analyzed by flow cytometry at different annual clinical visits to ascertain (**A**) the percentages of CD56^+^ NK cells among total lymphocytes and (**B**) ratio of CD16^+^CD56^dim^ to CD16^low^CD56^bright^ NK cell subsets. Gray bars represent the medians in each group. Each point represents data from a single individual. (**C-F**) NK cells from V2 or V3 for 4 non-sensitized (grey) and 4 sensitized (purple) patients were subjected to (**C**) UMAP dimensional analysis and (**D**) FlowSOM clustering. (**E**) Marker expression (MFI, mean fluorescence intensity) of 8 distinct subsets of NK cells within the UMAP distribution. (**F**) The proportional contribution (mean ± standard deviation) of NK cells from sensitized and non-sensitized children to each FlowSOM cluster. In **(A), (B)** and **(F)** *p* values were calculated using a Mann-Whitney U test. **p<0.05.

Despite these similar frequencies of NK cells, changes in the expression levels of germline encoded NK-cell receptors in different individuals could contribute to altered functional responses to inflammatory or environmental (e.g., allergen) cues. We used flow cytometry and dimensionality reduction analysis in the following manner: data visualization was performed by UMAP in conjunction with unsupervised clustering analysis by FlowSOM to project differences in NK-cell population phenotypes in a subset (described in **Table S3**) of allergen sensitized and non-sensitized children with AD based on the expression of CD16, CD56, NKG2D, CD69, CD62L, CCR4, Tim-3, NKp44, and CD45RO on NK cells (**Fig. 1C**). We chose these receptors based on evidence that their expression changes or contributes to NK-cell functional activity in allergic or asthmatic disease (*12, 20-24*). Eight distinct populations of NK cells were apparent in this FlowSOM analysis (**Fig. 1D+E** and **Fig. S2**). The CD16^dim^ CD56^bright^ NK cells cluster (#7, **Fig. 1D**) was similarly populated by cells from the sensitized and non-sensitized individuals (**Fig. 1F**), consistent with conventional flow cytometry analysis (**Fig. 1B**). Among the eight different NK-cell clusters, only two populations (Clusters 1 and 2) showed enrichment in non-sensitized compared to sensitized children (**Fig. 1F**). These two clusters were comprised of CD16^+^CD56^dim^ NK cells characterized by high expression levels of NKG2D, low expression of CCR4 and CD62L, and elevated expression of CD69 (**Fig. 1E)**. These results suggest that allergic sensitization in patients with AD is associated with alteration of the circulating NK cell repertoire.

### Expression of NKG2D on NK cells is inversely associated with severity of AD and with allergic comorbidities

The diminished presence of NKG2D^+^ NK cells in this subset of allergen-sensitized individuals (**Fig. 1**) may represent a specific early-life endotype of children with AD that reflects altered functional responsiveness to activating signals via NKG2D (*25*). Indeed, the reduced expression levels of NKG2D on CD56^dim^ NK cells in sensitized compared to non-sensitized MPAACH children (**Fig. 2A+B**) facilitated stratification and quantification of NKG2D^low^ and NKG2D^high^ NK cells (**Fig. S1C**). The phenotype of reduced NKG2D expression was specific to NK cells, as expression levels of NKG2D on non-NK cells (CD3^+^ or CD14^+^ or CD19^+^) did not differ between the sensitized and non-sensitized groups (**Fig. 2B**). The percentages of NKG2D^low^ CD56^dim^ (**Fig. 2C**) NK cells were increased in allergen-sensitized patients relative to non-sensitized children with AD at V2 (annual visit 2) and V3 (annual visit 3). CD56^bright^ NK cells showed a similar trend that did not reach statistical significance (**Fig. 2D**). The magnitude of difference in NKG2D expression levels was greatest at V3 (**Fig. 2C+D**, far right plots), suggesting a progressive aggravation of the NKG2D pathway.

**Figure 2.**
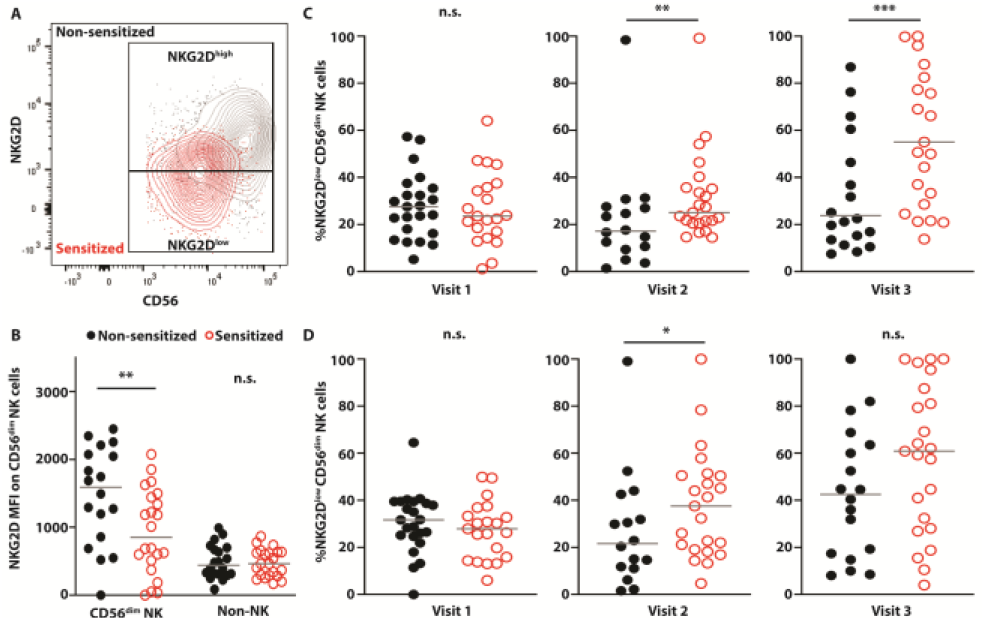
Progressive longitudinal accumulation of NKG2D^low^ CD56^dim^ NK cells in allergen sensitized children. (**A**) Representative plot of NKG2D^low^ and NKG2D^high^ proportions on CD56^dim^ NK cells determined by flow cytometry. (**B**) Mean fluorescence intensity (MFI) of NKG2D expression on CD56^dim^ NK cells and non-NK cells in non-sensitized (closed black circles, n = 16) and sensitized (open red circles, n = 23) children at V3. Proportion of (**C**) CD56^dim^ and (**D**) CD56^bright^ NK cells gated as NKG2D^low^ (defined in **A**) at V1, V2 and V3. Statistical differences between median values of non-sensitized (closed black circles, V1 n=23; V2, n=16; V3, n = 18) and sensitized (open red circles, V1, n =21; V2, n=23; V3, n =23) was determined by two-tailed Mann-Whitney U test. Gray bars represent the medians in each group. Each point represents data from a single individual: *p<0.09, **p<0.05, ***p<0.01.

Expression levels of the activating Fc receptor CD16 showed a similar progressive decline on the CD56^dim^ NK cell population in sensitized MPAACH children relative to those who are non-sensitized (**Fig. S3A**). There were no apparent trends or differences in the expression of CCR4, CD62L, and Tim-3 by CD56^dim^ NK cells between the two groups of children over time, while CD69 exhibited a consistent, non-significant reduction in expression on NK cells in sensitized compared to non-sensitized children (**Fig. S3B-D**). Similar non-significant trends were observed across the range of receptors analyzed in both CD56^bright^ (**Fig. S4**) and CD56^neg^ (**Fig. S5**) NK cells between sensitized and non-sensitized children.

Given the increasing trajectory of NKG2D^low^ NK cells over time in children with AD who had persistent or developed allergic sensitization, we examined the association of this NK-cell phenotype with the severity of AD. Sensitized children exhibited higher SCORAD (Scoring for Atopic Dermatitis, a clinical tool used to assess the extent and severity of eczema) than non-sensitized children (**Fig. S6**). Moreover, there was a positive association between the proportions of NKG2D^low^ CD56^dim^ NK cells and SCORAD in sensitized children that was not observed in non-sensitized children (**Fig. 3**). A similar trend was observed in CD56^bright^ NK cells (**Fig. S7**). Thus, reduced NKG2D expression on CD56^dim^ NK cells is linked to more severe eczema in allergen-sensitized children.

**Figure 3.**
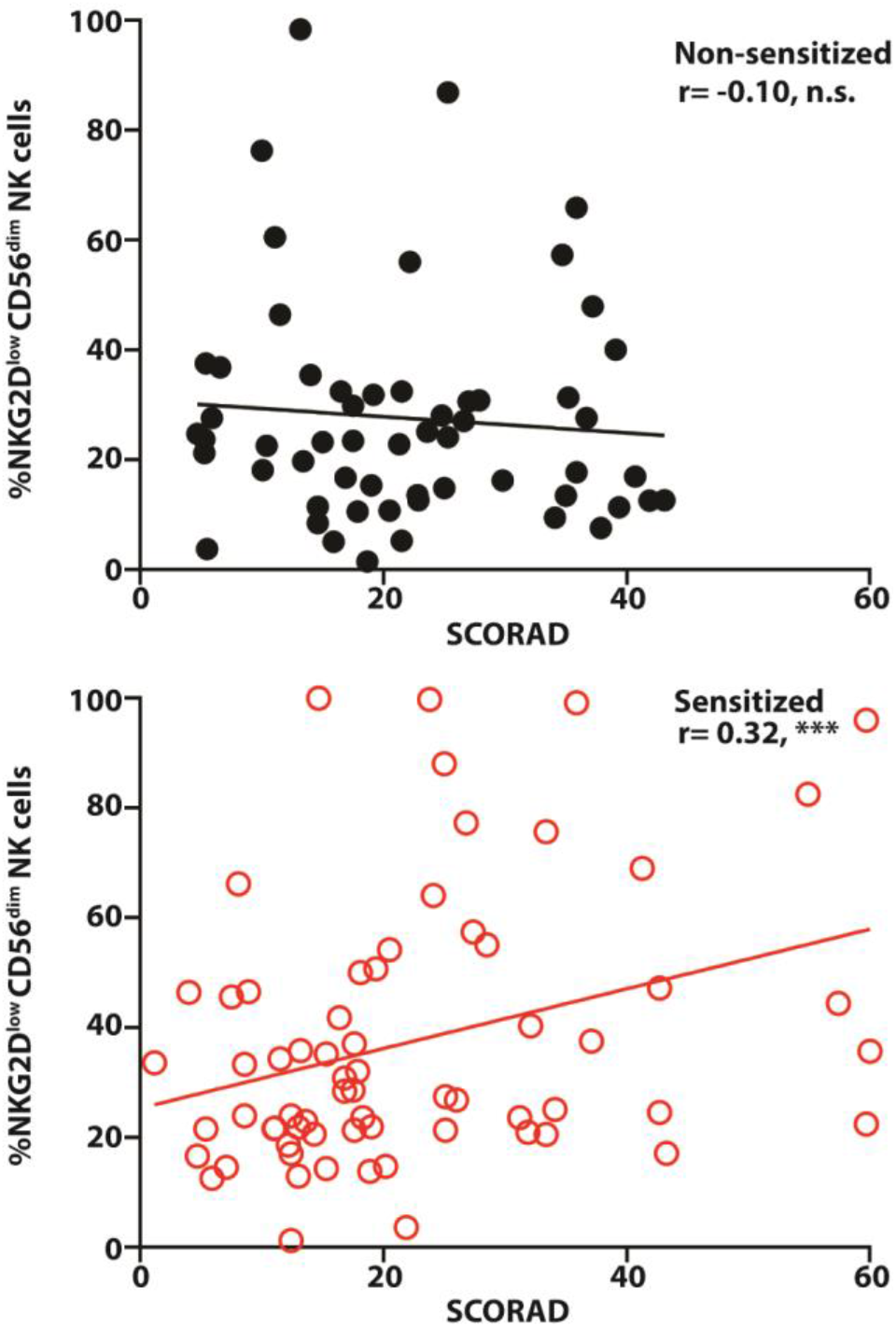
Association between NKG2D^low^ percentages and increased AD severity in sensitized children. Inverse relationship between SCORAD and proportion of NKG2D^low^ cells among CD56^dim^ NK cells was observed in sensitized (open red circles, n=67) but not in non-sensitized (closed black circles, n=57) children (Combined from V1-V3). Spearman correlation test was used to assess association between SCORAD and NK cells. Each point represents data from a single individual. *** p<0.01.

### NK cell phenotypic changes are most evident in children with co-sensitization to food and aero allergens

The type of allergic sensitization is important in both clinical outcome and treatment decision making. Specifically, co-sensitization to multiple allergens is a stronger risk factor for development of rhinitis and asthma compared to mono-sensitization (*26-28*). Since reduced NKG2D (**Fig. 2**) and CD16 (**Fig. S3A**) expression were characteristics of NK cell populations in AD patients with allergen sensitivity, we compared the expression of these receptors on NK cells in patients with sensitivity to one type of allergen (food or aero mono-sensitization) to patients with sensitization to at least one food and one aero allergen (co-sensitization). Similar to observations in **Fig. 2**, sensitization type had no effect on proportions of NKG2D^low^ CD56^dim^ NK cells at V1 (**Fig. 4A**, left column). An enrichment of NKG2D^low^ CD56^dim^ NK cells was observed at V2 and V3 in co-sensitized individuals but was not evident until V3 in mono-sensitized children (**Fig. 4A**, middle and right columns). A similar trend was evident for CD16 expression, with reduced expression levels of CD16 observed on the CD56^dim^ NK cell population in co-sensitized individuals at every clinical timepoint (**Fig. 4B**). This effect was not observed until V2 and was of a reduced magnitude in mono-sensitized children (**Fig. 4B**). Thus, co-sensitization is more strongly linked to reduced activating receptor expression on NK cells.

**Figure 4.**
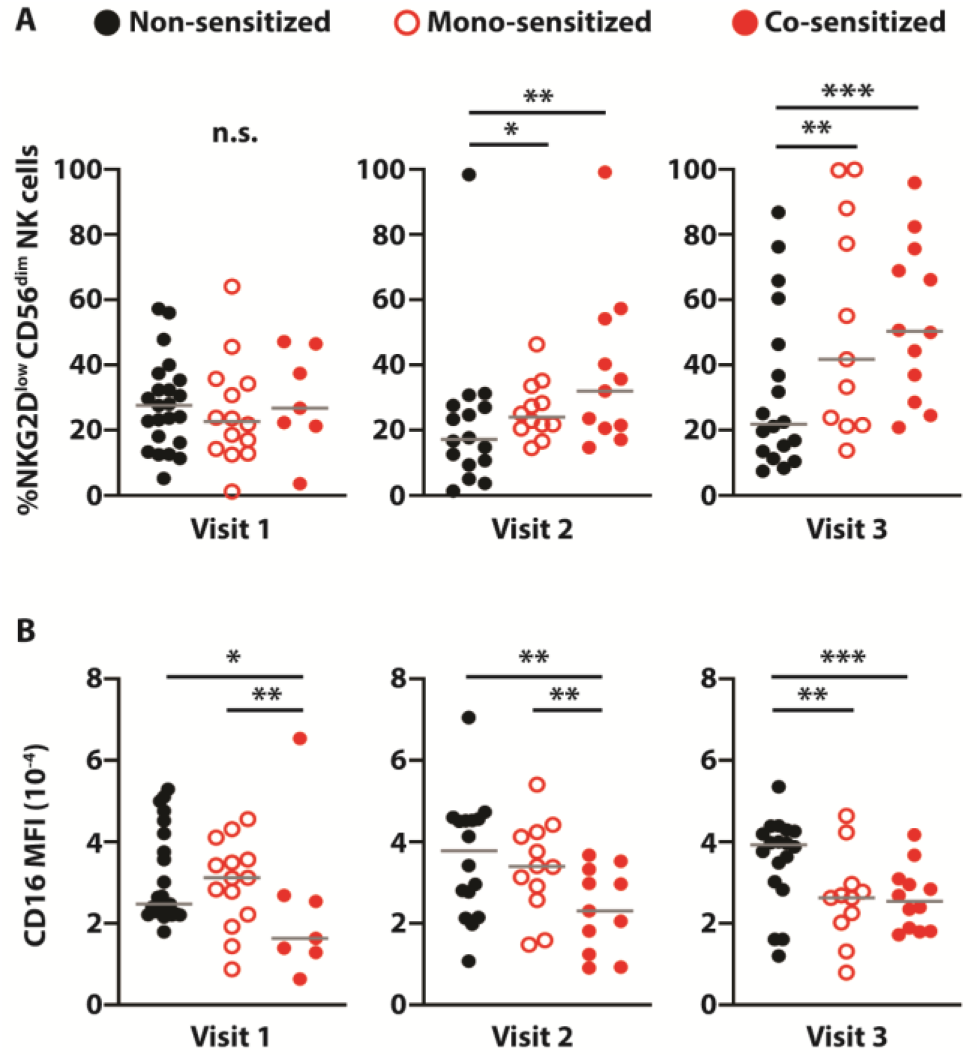
Accumulation of CD56^dim^ NK cells with reduced NK cell activating receptor expression is exacerbated by allergen co-sensitization. Flow cytometry was used to determine the proportion of (**A**) CD56^dim^ NK cells that are NKG2D^low^ or the (**B**) Mean fluorescence intensity of CD16 expression on CD56^dim^ NK cells at each clinical visit for children with AD. Children are separated into non-sensitized (closed black circles. V1, n=23, V2, n=16, V3, n = 18), sensitized to either food or aero allergens (monosensitization, open red circles. V1, n=14; V2, n=12; V3, n = 11), or sensitized to both food and aero allergens (co-sensitization, closed red circles. V1, n=7; V2, n=11; V3, n = 12) groups. Significant differences between group medians were determined by one-tailed Mann-Whitney U test based on expectation of singular direction of effect in Figure 2. Gray bars represent the medians in each group. Each point represents data from a single individual. *p<0.09, **p<0.05, ***p<0.01

### NK cells with reduced NKG2D expression display decreased cytolytic function

Numerous studies measuring cytolytic activity of peripheral NK cells against the K562 cells have shown that this activity is reduced in individuals with AD relative to healthy controls (*5-8*). Given the importance of NKG2D to NK cell killing of K562 cells (*29*), we evaluated whether cytolytic function would be more substantially impacted in sensitized individuals whose NK cells exhibit lower NKG2D expression. Prior to use in killing assays, PBMC were either unstimulated (**Fig. 5A**) or activated with IL-12 and IL-18 (**Fig. 5B**), which are established stimulants of NK cell activity (*30*) that are implicated in the pathogenesis of AD (*31-33*) and associated allergic diseases (*34-36*). Spontaneous killing activity observed in unstimulated NK cells from sensitized individuals was reduced compared to NK cells from non-sensitized children, which were similar to children without AD (non-AD, **Fig. 5A**). In addition, the capacity of IL-12 and IL-18 to amplify NK cell cytolytic activity was reduced more than two-fold in sensitized children relative to non-sensitized children with AD and non-AD controls (**Fig. 5B**). The CD56^dim^ NK cells from the sensitized individuals used in the functional assays depicted in **Fig. 5**, were confirmed to exhibit lower NKG2D expression compared to the CD56^dim^ NK cells from non-sensitized children used in the functional assays (**Fig. S8A**). While in this small subset it appeared that NKG2D was similarly decreased on CD56^dim^ NK cells from children without AD, this was not the case when we analyzed a larger whereby sensitized children overall showed the lowest levels of NKG2D expression relative to non-sensitized children and those children without AD subset (**Fig. S8B)**.

**Figure 5.**
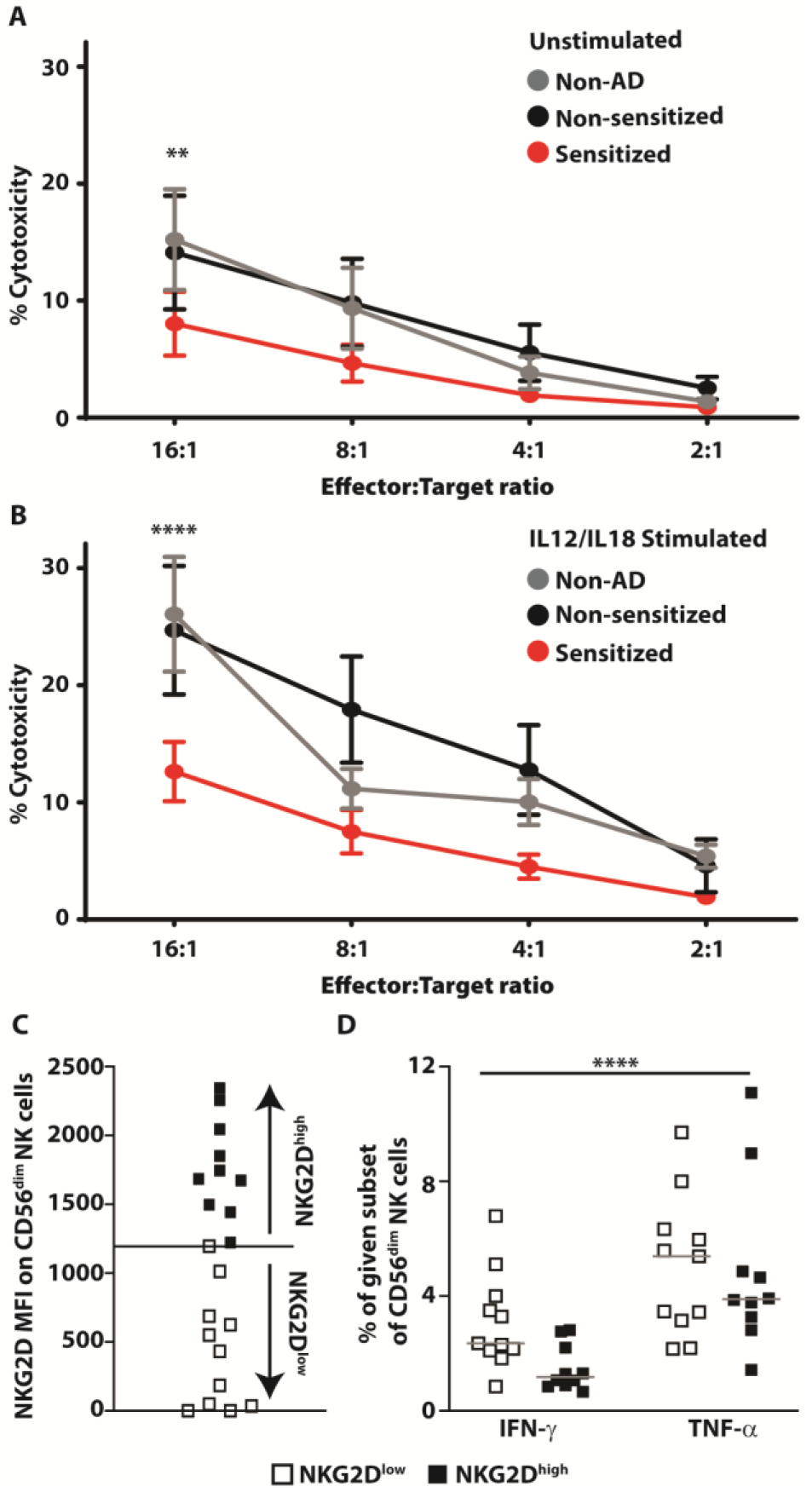
Reduced cytolytic function but enhanced NKG2D-induced cytokine production associated with low NKG2D expression. **(A-B)** Peripheral blood (V1 or V2) from children without AD (closed gray circles, n=4) or children with AD with allergen sensitivity (closed red circles,V1 or V2, n=5) or non-sensitized (closed black circles, V1 or V2, n=5) were stimulated with (**A**) media or (**B**) media containing IL-12 and IL-18 prior to 4-hour co-culture with ^51^Cr-labeled K562 cells. Chromium release into culture supernatant was measured using a gamma reader and determined as a proportion of max measured with detergent-medaited lysis of cells. Means and standard errors of each group were displayed. Comparison of the area under the curve was performed using ANOVA. (**C-D**) V3 Blood cells from n=21 patients were assigned into two groups based on (**C**) the median (black bar) of mean fluorescence intensity of NKG2D expression on CD56^dim^ NK cells and (**D**) Proportions of IFN-γ^+^ and TNF-α^+^ expression on gated subsets of NKG2D^low^ or NKG2D^high^ CD56^dim^ NK cells was measured by flow cytometry following stimulation with plate bound MICA (10 μg/ml). Mann-Whitney U test was used to determine significant difference between the groups with higher (closed black squares, n=10) and lower (open white squares, n=11) median NKG2D expression level. Gray bars represent the medians in each group. Each point represents data from a single individual. **p<0.05, ****p<0.001

### Peripheral NK cells from sensitized children with AD express higher levels of inflammatory cytokines

To assess the impact of reduced NKG2D expression on cytokine production by NK cells directly, we stimulated a subset of PBMC with plate-bound MICA, a known ligand for NKG2D (*25*). We utilized V3 samples (n=21) stratified as NKG2D^high^ or NKG2D^low^ (**Fig. 5C**) based on the observed expression level of NKG2D on NK cells by MFI (NKG2D^low^: below median; NKG2D^high^: above median, gated as shown in **Fig. S9A**). On average, patient-derived NK cells with lower than the median of NKG2D MFI responded to stimulation with MICA with greater cytokine production (IFN-γ and TNF-α) (**Fig. 5D**) than patient-derived NK cells with higher than the median of NKG2D MFI. In addition, blood CD56^dim^ NK cells from allergen sensitized children with AD at V1 and V2 (gated as shown in **Fig. S9B**) exhibited 45% greater spontaneous expression of TNF-α than non-sensitized children even in the absence of *ex vivo* restimulation (**Fig. 6A+B**). A similar trend for increased TNF-α expression was observed in CD56^bright^ NK cells (**Fig. 6C+D**). The expression of TNF-α remained elevated in these NK cell populations following stimulation with IL-12 and IL-18 (**Fig. 6B+D**). In contrast, IFN-γ expression by peripheral NK cells was only evident following stimulation with IL-12 and IL-18, and there were no significant differences observed in IFN-g expression levels between groups following this stimulation (**Fig. S10**). Similar to the overall cohort, there was a trend for lower NKG2D expression on NK cells from sensitized compared to non-sensitized children with AD in the subset of specimens used to measure inflammatory cytokine expression in **Fig. 6** (**Fig. S11**). Consistent with progressive aggravation of NKG2D expression at later clinical visits (**Fig. 2**), the MFI of NKG2D expression was inversely correlated with TNF-α production in samples derived from V2 but not those from V1 (**Fig. S12**).

**Figure 6.**
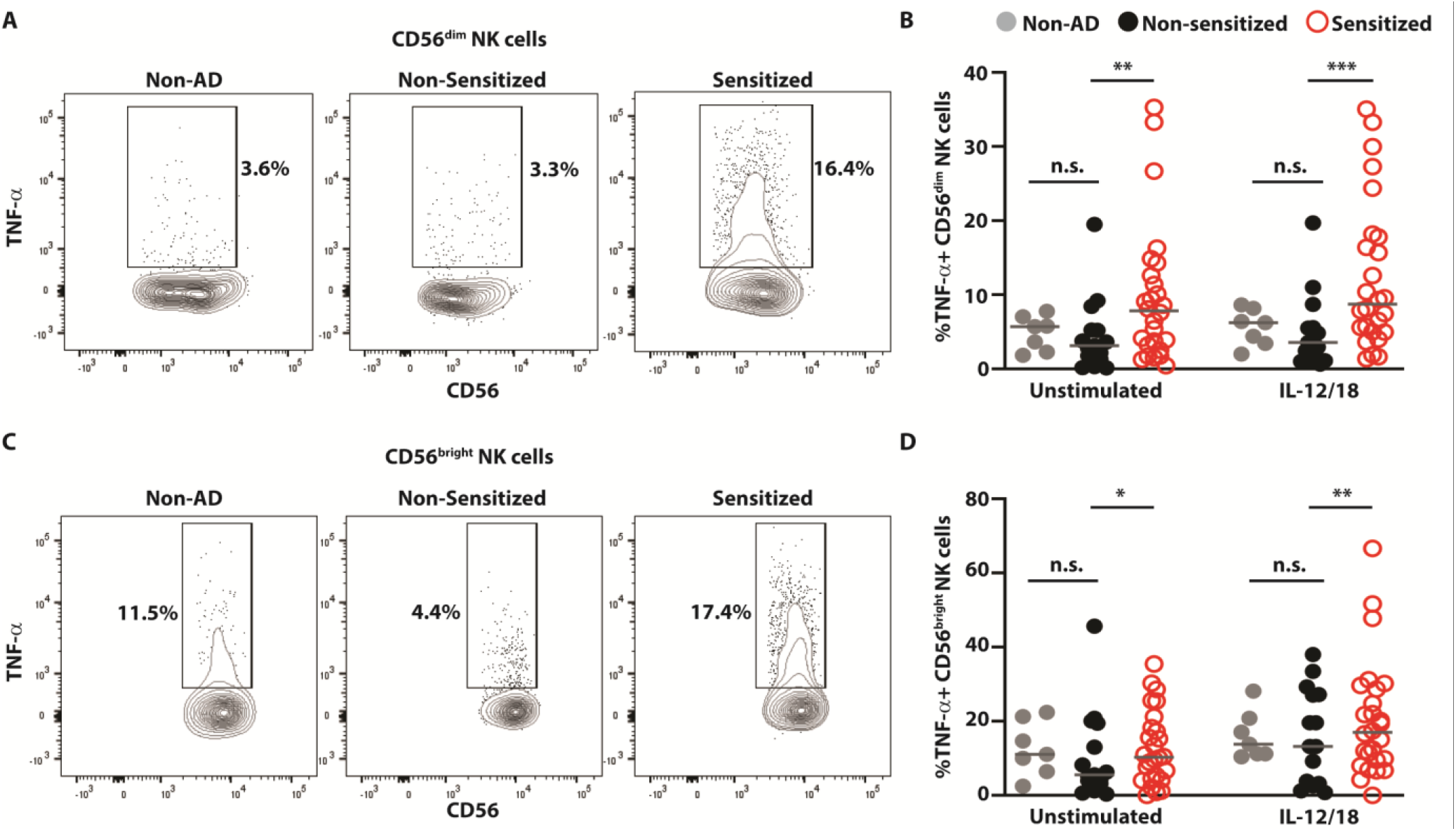
CD56^dim^ and CD56^bright^ NK cells in allergen-sensitized patients show increased expression of TNF-α. Percent TNFα^+^ (**A-B**) CD56^dim^ and (**C-D**) CD56^bright^ NK cells following culture of V1 and V2 peripheral blood cells in media in the absence or presence of IL-12 and IL-18. NK cells derived from PBMC from MPAACH children who were: non-AD (closed gray circles, n = 7), non-sensitized (closed black circles, n=15), or allergen sensitized (open red circles, n=26) were evaluated for cytokine expression by flow cytometry. Statistical differences between group medians were determined by one-tailed Mann-Whitney U test. Gray bars represent the medians in each group. Each point represents data from a single individual. *p<0.09, **p<0.05, ***p<0.01

### Individual-level longitudinal analysis of NKG2D^low^CD56^dim^ and NKG2D^low^CD56^bright^ NK cells reveals their association with persistent and acquired sensitization

In a subset of 10 children for whom individual-level longitudinal NK cell and sensitization data was available across multiple time points, we examined NK cells and allergic sensitization (non/transient vs. acquired/persistent) within each child over time. There were no differences in age, sex, race or public insurance in those who were non/transiently sensitized compared to those that were acquired/persistently sensitized (**Table S4)**. At V3, children with acquired or persistent sensitization had increased proportions of NKG2D^low^CD56^dim^ cells (**Fig. S13** and **Fig. 7A**) compared to children who were non-sensitized or had transient sensitization. A similar trend was observed in proportions of NKG2D^low^CD56^bright^ NK cells (**Fig. 7B**). Over time from V1/2 to V3, children with acquired/persistent sensitization had a mean increase of 28.6% in NKG2D^low^CD56^dim^ cells and 29.6% in NKG2D^low^CD56^bright^ NK cells compared to only 2.7% and 2.9% for the non/transiently sensitized children (**Fig. 7 C+D**). We next evaluated whether the change in the percentage of NK cells over time from V1/V2 to V3 was associated with AD severity and/or skin barrier function assessed by transepidermal water loss (TEWL). Lesional TEWL was positively associated with an increase in NKG2D^low^ NK cells for both the CD56^dim^ (**Fig. S14A**) and CD56^bright^ (**Fig. S14B**) subsets. When we stratified by sensitization phenotype, we observed that these positive associations were driven by the children with longitudinal acquired/persistent sensitization (red lines in **Fig. S14 C+D**) rather than non/transient sensitization (black lines). There were no associations between NK cell proportion over time and SCORAD (**Fig. S14E-H**), regardless of stratification. Collectively, these data suggest that the progressive enrichment in NKG2D^low^ NK cells over time is co-incident with worsening skin barrier function and persistent or late sensitization.

**Figure 7.**
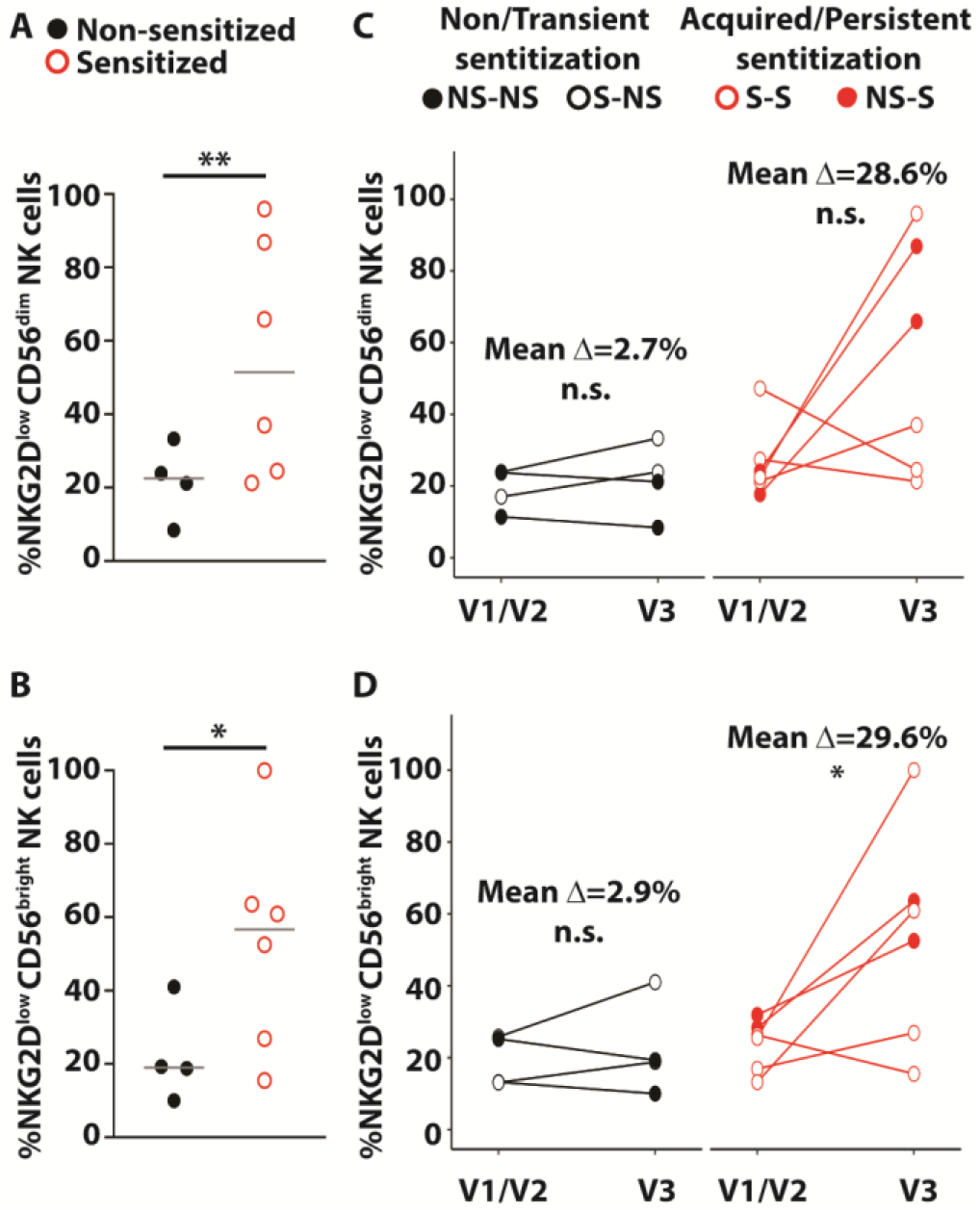
Individual-level longitudinal analysis of proportions of NKG2D^low^ CD56^dim^ and CD56^bright^ NK cells in MPAACH participants with different longitudinal sensitization patterns as indicated. CD56^dim^ and CD56^bright^ NK cells NKG2D^low^ measurements in participants at V1/V2 and V3 were plotted individually to provide summary of non-(NS-NS) or transient (S-NS) sensitized (closed black circles, n = 4) vs persistent (S-S) or acquired sensitized (NS-S) (open red circles, n = 6) at V3 for (**A**) CD56^dim^ and (**B**) CD56^bright^ NK cells. Trajectories of proportions of (**C**) NKG2D^low^CD56^dim^ and (**D**) NKG2D^low^ CD56^bright^ NK cells within individual children over time who have the following longitudinal sensitization patterns: non-sensitization (NS-NS, closed black circles, n = 2), transient sensitization (S-NS, open black circles, n = 2), acquired sensitization (NS-S, closed red circles, n = 2) and persistent sensitization (S-S, open red circles, n = 4). In **(A)** and **(B)** significant differences between group medians were determined by one-tailed Mann-Whitney U test based on expectation of singular direction of effect in Figure 2. In **(C)** and **(D)**, significance was evaluated using a paired t-test. Gray bars represent the medians in each group. Each point represents data from a single individual *p<0.09, **p<0.05.

## Discussion

Numerous publications over the past four decades have compared patients with AD to healthy controls and established a paradigm that loss of NK-cell activity contributes to pathogenesis of eczema (*10-15*). As a result, several translational studies are focused on therapeutic expansion or restoration of NK cells in patients with AD. Our analyses in a well-defined longitudinal cohort of children with AD surprisingly revealed a progressive accumulation of hyperinflammatory NKG2D^low^ NK cells that is associated with more severe AD and allergen sensitivity. The loss of NKG2D expression by NK cells was more pronounced in patients co-sensitized to both aero and food allergens, a known risk factor for development of asthma. An individual-level analysis of longitudinal paired sensitization and NK cell data over time further supported these findings. Strikingly, there was a marked increase in the proportion of NKG2D^low^ NK cells over time in children who were persistently sensitized or acquired sensitization over time in contrast to those who were non-sensitized or only transiently sensitized early. Further, this magnitude of the increase in NKG2D^low^ NK cells was correlated with skin barrier dysfunction in these individuals. Collectively, our data indicate that dysfunctional hyperinflammatory NKG2D^low^ NK cells accumulate co-incidentally with worsening sensitization and increasing barrier dysfunction. These observations provide important insights into a potential pathophysiological mechanism of atopic march involving altered NK cell functional responses.

Atopic dermatitis and associated allergic comorbidities are correlated with heightened expression of type 2 cytokines, including IL-4 (*37*). Therefore, one potential mechanism of reduced NKG2D expression and diminished cytolytic function of NK cells relates to potential effects of the type 2 cytokines on NK cells. Indeed, IL-4 is capable of down-regulating NKG2D expression on NK cells (*38*). In contrast, type 1 cytokines like IL-12 can promote increased expression of NKG2D (*39*), although this may be counteracted by suppressive effects of IL-4 on IL-12 responses (*40*). A potential source of type 2 cytokines are T cells (*41*). In a recent study by Zhou et al., dysregulation of NK-cell surface markers expression and function was shown to be downstream of Th2 allergic T cells responses (*42*). Whether other type 2 cytokines such as IL-13, IL-5, IL-33, and TSLP can influence expression of NKG2D remains to be determined.

A second mechanism potentially contributing to downregulation of NKG2D on NK cells in allergen sensitized patients with AD lies in the effects of allergens themselves. Children sensitized to house dust mite exhibit increased sera levels of soluble ligands (e.g. MICA and ULBP2) for NKG2D compared to non-sensitized children (*43*). One effect of elevated expression of soluble NKG2D ligands is the downregulation of NKG2D expression on NK cells (*44, 45*). Thus, reduced NKG2D expression in allergen sensitized patients might be caused by allergen-induced increases in the levels of circulating soluble NKG2D ligands. Allergens often also possess proteolytic activity. Since metalloproteases like ADAM10 play a role in cleavage of both NKG2D ligands and the receptor CD16 from the surface of NK cells (*46, 47*), similar functionality of allergen-associated proteases or proteolytic activation of endogenous metalloproteinases may contribute to loss of NKG2D and CD16 expression. These observations may also explain the exacerbation of NKG2D loss in co-sensitized patients, who respond to a greater number of potential allergens and increased frequency of allergen exposure to amplify these effects. Moreover, aero allergens are more difficult to effectively avoid compared to food allergens and might favor chronic stimulation of these putative pathways influencing NK cell phenotype and function.

NK cells sense stressed cells via NKG2D to detect a repertoire of surface expressed stress ligands **(*48, 49*)**. Oncologic transformation, microbial infections or environmental stressors promotes the expression of NKG2D ligands (*49-51*). Here we show that NKG2D interaction with its ligands promotes NK cells activation **(Fig. 5)**. Moreover, the above-described interactions allow NK cells to eliminate virally transfected cells and stimulate myeloid cells (*52, 53*). Thus, we can speculate that reduced expression of NKG2D would be reflected in reduced ability to combat pathogens. Several studies showed increased susceptibility to viral and bacterial infections in children with AD (*54, 55*). Recently, a study conducted by Medeleanu et al. suggested that poly-sensitized children are more susceptible to lower respiratory tract infection in early life in addition to an increased association to AD comorbidities (*56*). Thus, our data supports the idea that NKG2D and CD16 are downstream to allergen sensitization, which potentially affects antimicrobial response in AD.

One curious observation in our report is the positive relationship between the severity of AD and the proportion of NKG2D^low^ NK cells in sensitized children. Our analysis, as well as other studies, showed an association between allergen sensitization and breach in the skin barrier (*57*). Thus, interpretation of these data suggests that reduced integrity of skin barriers combined with sensitization may contribute to the downregulation of NKG2D. This may relate to internalization of allergens or inflammatory stimulants through breaches in the epidermal barrier. Indeed, introduction of *Staphylococcus aureus* through the skin barrier at sites of eczema lesions would provide additional ligands for NKG2D on monocytes (*58*) that could subsequently stimulate NK cells and promote loss of NKG2D. A second interpretation of our data holds that accumulation of hyperinflammatory NKG2D^low^ NK cells in certain patients could exacerbate epidermal integrity disruption resulting in increased transdermal exposure to allergens. This hypothesis is supported by the progressive decline in NKG2D expression by NK cells over subsequent clinical visits.

TNF-α impairs skin barrier function (*59, 60*) and could therefore contribute to worsening of AD as well as increased translocation of potential allergens. Of note, elevated TNF-α expression by NK cells is also a characteristic of severe asthmatic patients (*22*). The increased capacity for IFN-γ and TNF-α secretion by NKG2D^low^ NK cells present in sensitized patients in response to MICA may also contribute to the pathophysiology of atopic march. Indeed, an IFN-γ signature in the context of elevated type 2 cytokines is a characteristic of severe asthma (*22, 23, 61, 62*). In addition, activated NK cells can produce cytokines including TNF-α *(16)* and GM-CSF *(17)* that may aggravate allergic disease mechanisms (*18-21*). Indeed, disease pathogenesis may depend on a feedback loop between skin barrier disruptions (*63*) affecting NK-cell functionality that in turn exacerbates skin barrier disruption to amplify allergen exposure (*64*).

Reduced NKG2D- and CD16-mediated cytolytic functionality of NK cells in allergen-sensitized patients with AD may also undermine key immunoregulatory mechanisms that limit development of allergy, allergic rhinitis, and asthma. NK cell killing of dendritic cells (*10*), CD4 T cells (*65*), and eosinophils (*66*) is likely to limit development of allergic disease. Therefore, the diminished capacity of NKG2D^low^ NK cells in allergen sensitized patients with AD may prime the development of more severe allergic diseases and asthma. Of note, the development of asthma is typically established early in life and has been shown to be associated with increased microbial infections (*67, 68*).

Our study has several limitations. The individual-level longitudinal analysis was performed on a relatively small number of samples. This prevented us from achieving additional conclusions regarding the association of reduced NKG2D expression with other AD clinical outcomes such as food sensitizaiton, food allergy or allergic rhinitis. Moreover, the constrained volumes of patient blood accessible for this sub-study and limited availability of optimized high-dimensional flow cytometry panels resulted in a rather narrow phenotypic analysis of NK cells. An expanded panel of markers or unbiased approaches (i.e. single-cell RNA sequencing) likely would have provided deeper insights into both phenotypic and functional changes in subsets of NK cells within this patient population. In addition, the data described in the current manuscript is limited to analysis of circulating blood NK cells. Since AD is a skin disease with allergic manifestations related to respiratory and intestinal mucosa, additional insights would likely be gained from analysis of NK cells in the skin, gut, and lungs. However, since MPAACH is a cohort of young children, it was not feasible to obtain NK cells from any of the above-mentioned tissues. Finally, despite longitudinal evidence for progressive exacerbation of the observed phenomena and relationship to eczema severity, our study could not authoritatively elucidate a pathological role for NKG2D^low^ NK cells in AD sequelae.

Overall, our data show that a progressive accumulation of hyperinflammatory NKG2D^low^ NK cells defines an endotype of severe eczema associated with allergen sensitivity. These observations reveal a pathophysiological mechanism of NK cell dysregulation in AD that potentially contributes to atopic march. Therefore, the current paradigm of NK-cell deficiency in AD and associated translational focus on therapeutic restoration of NK cell activity may need to be re-evaluated in light of possible pathogenic contributions of NK-cell cytokine production to skin barrier disruption and development of allergic comorbidities in this patient population.

## Materials and Methods

### Study design

We characterized peripheral NK cells over time in children with AD participating in the MPAACH cohort. We defined phenotypic and functional changes in the peripheral NK cells and related these to sensitization, AD severity, and skin barrier function. For a subset of children for whom individual-level longitudinal NK cell data was available, analysis of the proportions of NKG2D^low^CD56^dim^ and NKG2D^low^CD56^bright^ NK cells expression was analyzed across multiple visits within the same child to determine their trajectories. The longitudinal analysis of NKG2D measurements on CD56^dim^ and CD56^bright^ NK cells was performed for each individual child within each sensitization phenotype.

### Study Subjects

MPAACH is a prospective longitudinal early life cohort of children with AD that has been previously described (*18*). Recruitment began in December of 2016 and is ongoing. Briefly, children in the greater Cincinnati area (southwest Ohio and northern Kentucky) were identified through public advertising or the medical chart at Cincinnati Children’s Hospital Medical Center.

Eligible children were aged ≤2 years upon enrollment, had a gestation of ≥36 weeks and either a diagnosis of AD (based on the Hanifin and Rajka Criteria for Atopic Dermatitis (*69*)) or the parent(s)/legal authorized representative indicated a positive response to each of the 3 questions from the Children’s Eczema Questionnaire (CEQ) (*70*). Each year following enrollment, MPAACH children undergo annual visits during which clinical data is obtained through questionnaires, extensive biospecimens are collected, and the children undergo skin prick testing (described below). At age 7, pulmonary function testing is performed. In a subset of experiments, children without AD were also recruited to serve as controls for baseline NK-cell functional responses. A set of adult peripheral blood mononuclear cells (PBMC) were used as a phenotyping control. This study was approved by the Institutional Review Board at Cincinnati Children’s Hospital Medical Center, and all subjects signed informed consent/parental permission prior to participation. Approved IRB 2016-5842.

### Clinical Outcome Definitions

At each annual visit, MPAACH children undergo assessment of the severity of their AD using Scoring for Atopic Dermatitis (SCORAD), a clinical tool used to assess the extent and severity of eczema based on the area affected and the intensity of redness, swelling, oozing/crusting, scratch marks, skin thickening and dryness (*63*). The quality of each child’s skin barrier function is assessed by quantification of transepidermal water loss (TEWL) at both lesional and never-lesional sites at each visit using the DermaLab TEWL probe (Cortex Technology, Hadsund, Denmark) as previously described (*18*). All children undergo annual skin prick testing (SPT) to 11 aeroallergens and 6 foods. The aeroallergen panel includes mold mix 1 (*Alternaria tenuis, Hormodendrum Cladosporioides, Helminthosporium interseminatum, Aspergillus fumigatus, Aspergillus Niger, Penicillium notatum*), mold mix 2 (*Rhizopus nigricans, Pullularia pullulans, Fusarium vasinfectum, Mucor racemosus*), ragweed (*Ambrosia trifida, Ambrosia artemisiifolia*), grass (Kentucky Bluegrass, Orchard, Redtop, Timothy, Sweet Vernon), tree mix 1 (Pecan, Maple BHR, Oak RVW, American Sycamore, Black Willow), tree mix 2 (White Ash, Birch Mix (Red&White), Black Walnut, Common Cottonwood, & American Elm), weeds (Kochia, English Plantain, Lamb’s Quarters, Marshelder/Poverty (BPT Mix) Common Cocklebur, & Careless/Pigweed (CR Mix)), mite mix (*Dermatophagoides pteronyssinus* and *D. farina*), cockroach (*Periplaneta americana* and *Blattella germanica*), cat and dog (HollisterStier, Spokane, WA). The food allergen panel includes cow’s milk, egg white, egg yolk, soy, wheat, tree nuts (almond, brazil nut, cashew, hazel nut, pecan, pistachio, and walnut), and peanut (Greer, Lenoir, NC). A positive SPT is defined as a wheal measurement 3mm or larger than the diluent control. Co-sensitization was defined as a positive SPT to at least one or more aeroallergens and one or more foods. Four longitudinal individual-level sensitization phenotypes were defined based on sensitization data from V1, V2 and V3 in the same child: *persistently non-sensitized* (non-sensitized at V1, V2 and V3); *transient* (sensitized at V1 or V2, but not at V3); *acquired* (non-sensitized at V1 and/or V2 but sensitized at V3); and *persistently sensitized* (sensitized at V1 and/or V2 and V3).

### Blood Sample Collection and Processing

Whole blood samples collected from MPAACH children were used to isolate human PBMC using Ficoll®Paque Plus (Sigma-Aldrich, Inc. St. Louis). The concentration of white blood cells in PBMC was measured using DxH 520 Hematology Analyzer (Beckman Coulter, CA). PBMC were resuspended in 1 mL of RPMI 1640 medium with 20 % FBS and stored at 4°C for immediate analysis or at -80°C for future studies.

### Antibodies

The following conjugated antibodies (Abs) were used in the described studies: Brilliant violet (BV) 421 anti-CD56 (5.1H11), BV711 anti-CD16 (3G8), Alexa-Fluor (AF) 647 anti-NKp44 (9E2), PE anti-Tim-3 (P44-8), PE/Cy7 anti-CCR4, APC anti-IFN-γ (4S.B3), PE anti-TNF-α(MAb11), PE/Cy7 anti-CD107a (H4A3) and AF488 anti-CD45RO were purchased from Biolegend (San Diego, CA); Brilliant Ultra violet (BUV) 395 anti-CD3 (SK7), V500 anti-CD3 (SP34-2), V500 anti-CD19, V500 anti-CD14, BUV737 anti-CD69 (FN50), and BUV395 anti-CD62L (SK11) were purchased from Becton and Dickinson (BD, San Diego, CA); eFluor710 anti-NKG2D (1D11) from Invitrogen (Carlsbad, CA); Abs were used at doses titrated in our lab.

### Sample analysis and internal controls

All phenotypic analyses were performed on fresh samples directly *ex vivo* to avoid complications of freeze/thaw. To control for batch effects across the cohort, we: (1) used the same antibody clones (as mentioned above), flow cytometer (LSR Fortessa instrument, BD Biosciences), staining protocol and reagents: Brilliant Violet staining buffer (BD Biosciences), running buffer (MACSQuant®, Miltenyi Biotec, Bergisch Gladbach, Germany) and fixation buffer (eBioscience™ IC Fixation Buffer, Invitrogen) through the entire study, (2) samples analysis was performed in weekly batches post sample fixation, and (3) included an internal control of healthy adult PBMC cultured with IL-2 (200U/mL) per each experimental batch (week). The internal controls were used as a benchmark for the manual gating of the samples obtained as part of the MPAACH cohort.

### NK-cell activation assays

Thawed PBMC were cultured at 2×10^6^ cells per well in the presence of 200 U/mL of human IL-2 (Peprotech®, Rocky Hill, NJ) for 18 hours. After 18 hours, incubated cells were stimulated in the presence or absence of plate bound major histocompatibility complex class I chain-related A (MICA) (10 μg/mL) for 6 hours. Golgistop™, GolgiPlug™ (BD), and PE/Cy7 anti-CD107a antibody (1:200) were added for the final five hours of the incubation (according to manufacturer guidelines). At harvest, cells were stained for surface markers before intracellular cytokines by utilizing eBioscience™ Intracellular Fixation & Permeabilization buffer set (Invitrogen). Flow cytometric data for phenotypic and functional analysis was obtained using an LSR Fortessa instrument and analyzed via FlowJo_v10.8.1 software (Treestar). Samples were selected for NKG2D functional assays from MPAACH children at V3 based on NKG2D-MFI expression.

### NK-cell cytokine production assessment

Thawed PBMC were cultured at 2×10^6^ cells per well in the presence of 200 U/mL of human IL-2 (Peprotech®, Rocky Hill, NJ) for 18 hours. After 18 hours of incubation, cells were stimulated in the presence of IL-12 and IL-18 (2.5 ng/mL and 10 ng/mL) or media for 36 hours. At the last five hours of incubation, Golgistop™ and GolgiPlug™ (BD) were added (according to manufacturer’s guidelines). Cells were harvested and stained for surface markers and then for the expression of intracellular cytokines by utilizing eBioscience™ Intracellular Fixation & Permeabilization buffer set (Invitrogen). Flow cytometric data for all phenotypic and functional analysis were obtained using an LSR Fortessa instrument (BD Biosciences) and analyzed via FlowJo_v10.8.1 software (Treestar).

### Flow cytometry dimensionality reduction and unsupervised cluster analysis

The visualization of dimensionality reduction analysis was performed by utilizing Flowjo packages (Down sample, concatenation, Uniform Manifold Approximation and Projection (UMAP) (*71*) and unsupervised clustering was performed via FlowSOM, we instructed FlowSOM to generate 8 clusters of NK cells (*72*). Only samples containing at least 10,000 gated NK cells were utilized to ensure sufficient numbers of cells for generation of the concatenated file after downsampling (10,000 NK cells/file) for statistically robust downstream analyses by UMAP and FlowSOM.

### NK-cell cytotoxicity assay

K562 target cells were labeled for 2 hours with 2μCi of sodium chromate (Na_2_^51^CrO_4_) per 2 × 10^4^ target cells at 37 °C, 5% CO_2_ and then washed to remove excess ^51^Cr. Labeled target cells were added to 96-well round-bottom plates (1 × 10^4^ cells/well). Isolated NK cells were added to the plates with E:T ratios ranging between 16:1 and 1:1. The amount of ^51^Cr released, which corresponds to target cell death, was measured by a gamma scintillation counter. The percent cytotoxicity against target cells was calculated with the following formula: %Cytotoxicity= 100×((experimental lysis – spontaneous lysis)/ (maximal lysis – spontaneous lysis)). To determine maximal lysis, ^51^Cr-labeled target cells were treated with 3% Triton X for 4 hours. To determine spontaneous release, target cells incubated without effector cells were used.

### Statistical analyses

Statistical analyses were performed in GraphPad Prism 9.5 (GraphPad software, CA) and R version 4.1.0 (R core team 2021). Prior to the analysis, Shapiro-Wilk tests were performed to examine the distribution of the data for each continuous variable. Descriptive statistics are presented in median (25^th^-75^th^ percentile) or frequency (%) due to the non-normal distributions. Comparisons between the full cohort and the subset were conducted using Mann-Whitney U and Fisher’s exact tests for continuous and categorical variables, respectively. The frequency of each NK-cell subset was compared using Kruskal-Wallis test across visits. Spearman correlation coefficients were used to measure the strength of association between NKG2D and SCORAD and TEWL.

## Data Availability

All data produced in the present work are contained in the manuscript and raw data will be made available upon publication or reasonable request to the authors

## Acknowledgements

We thank the participating children and families and all of the staff involved with the MPAACH study.

## Funding

This work was supported by 5U19AI70235 (GKKH, JBM, LJM); American Heart Association, 5T32ES010957 (DEO); AR073228 (SNW); 5U19AI070535 (SA); and 5T32GM063483 (SBD).

## Author contributions

D.E.O. contributed to the design of the study, performed experiments, interpreted results, acquired funding for the project, drafted the manuscript and reviewed and approved the final manuscript as submitted. S.B.D. performed experiments and reviewed and approved the final manuscript as submitted. W.C.C. and S.A. performed data analyses, interpreted results and reviewed and approved the final manuscript as submitted. D.K. and H.S. performed experiments and reviewed and approved the final manuscript as submitted. B.G. and D.S. contributed to the acquisition of data and reviewed and approved the final manuscript as submitted. L.J.M. and J.M.B. interpreted results and reviewed and approved the final manuscript as submitted. S.N.W. and G.K.K.H. contributed to the conception and design of the study, oversaw data analyses, interpreted results, acquired funding for the study, drafted the manuscript and reviewed and approved the final manuscript as submitted.

## Data and materials availability

All data needed to evaluate the conclusions in the paper are available in the main text or the Supplementary Materials.

## SUPPLEMENTAL MATERIAL

**Fig. S1.**
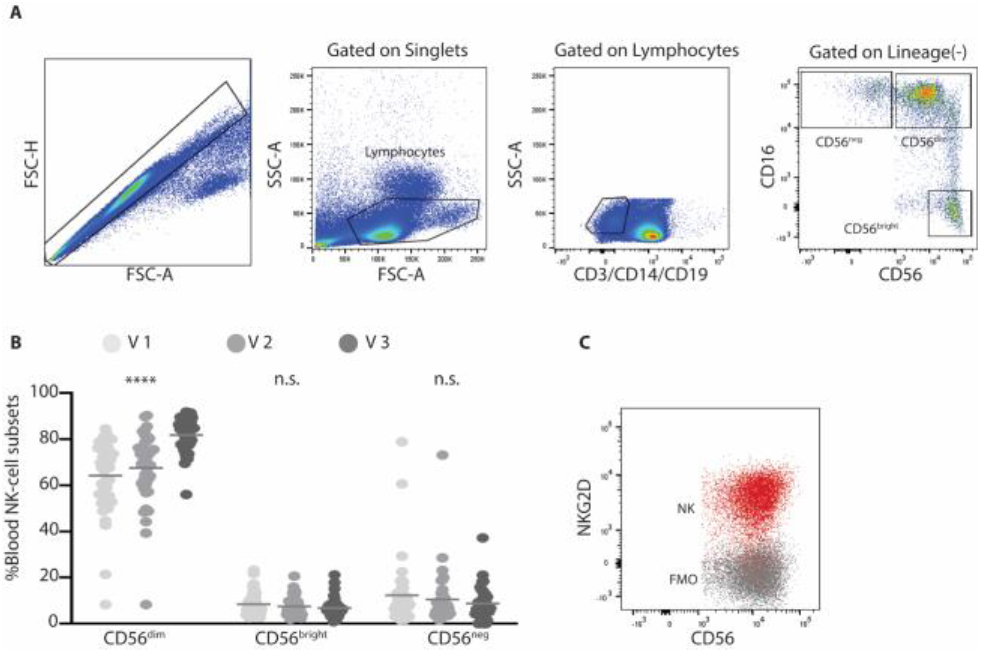
Age-dependent changes in blood NK cells subset composition in children in the MPAACH cohort. **(A)** Representative flow cytometry analysis with NK cells sequentially gated as singlet, live lymphocyte, and lineage negative (CD3^neg^ CD19^neg^ CD14^neg^) events prior to discrimination of CD16^+^CD56^neg^, CD16^+^CD56^dim^, or CD16^neg^ CD56^bright^ NK cells. (**B**) Median values of the frequency of each NK-cell subsets at each of three annual study visits, V1 (light grey circles, n=44), V2 (grey circles, n=39), V3 (dark grey circles, n=41). Gray bars represent the medians in each group. Each point represents data from a single individual. Comparison of medians was done using Kruskal-Wallis test. ****p<0.001. (**C**) Strategies to gate NKG2D in NK cells relative to IL-2-treated positive control NK cells (red) and FMO control (dark).

**Fig. S2.**
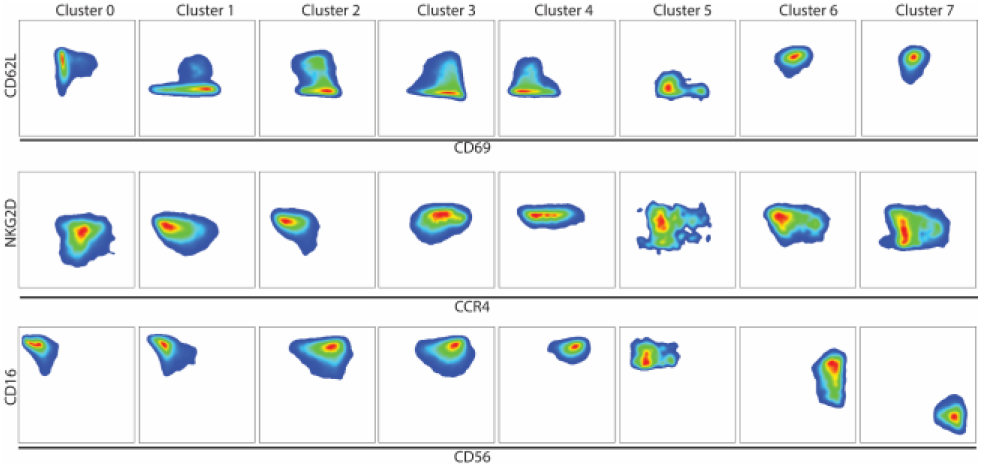
Characterization of Uniform Manifold Approximation and Projection clusters. Analysis of surface marker expression on the various clusters of NK cells presented in Figure 1E.

**Fig. S3.**
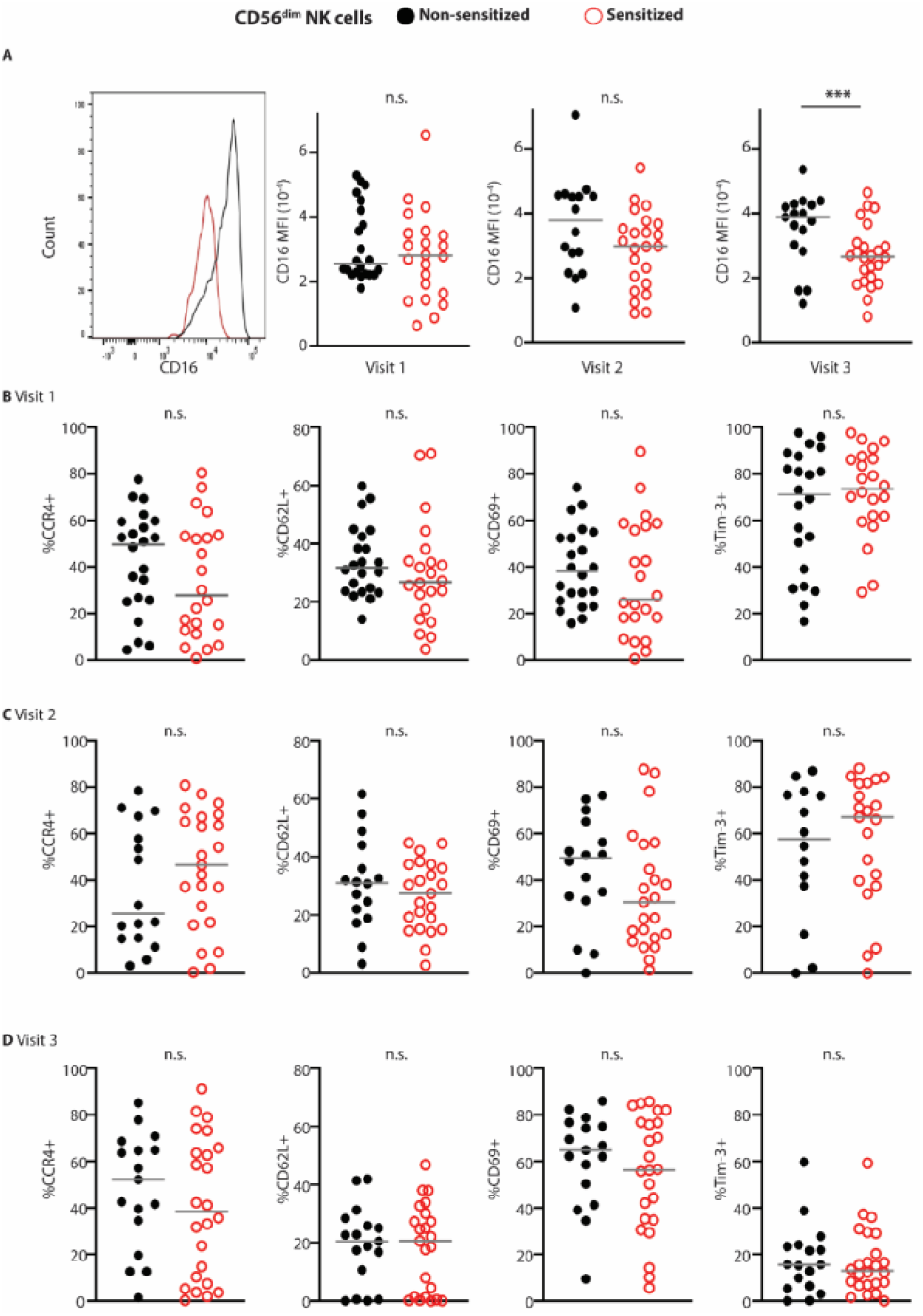
Allergen sensitization dependent changes in CD56^dim^ NK cell receptor expression across three annual visits. The expression of various surface markers on non-sensitized (closed black circles, V1, n=23; V2, n=16; V3, n=18) and sensitized (open red circles, V1, n=21; V2, n=23; V3, n=23) (**A**) Representative histograms of mean fluorescence intensity showing reduced CD16 expression on NK cells from sensitized patients at V3 and comparison between medians of CD16 per visit. (**B-D**) Proportions of CD56^dim^ NK cells in each group of patients expressing CCR4, CD62L, CD69, and Tim-3 at V1-V3. Gray bars represent the medians in each group. Each point represents data from a single individual. Significant differences between group medians were determined by Mann-Whitney U test. ***p<0.01.

**Fig. S4.**
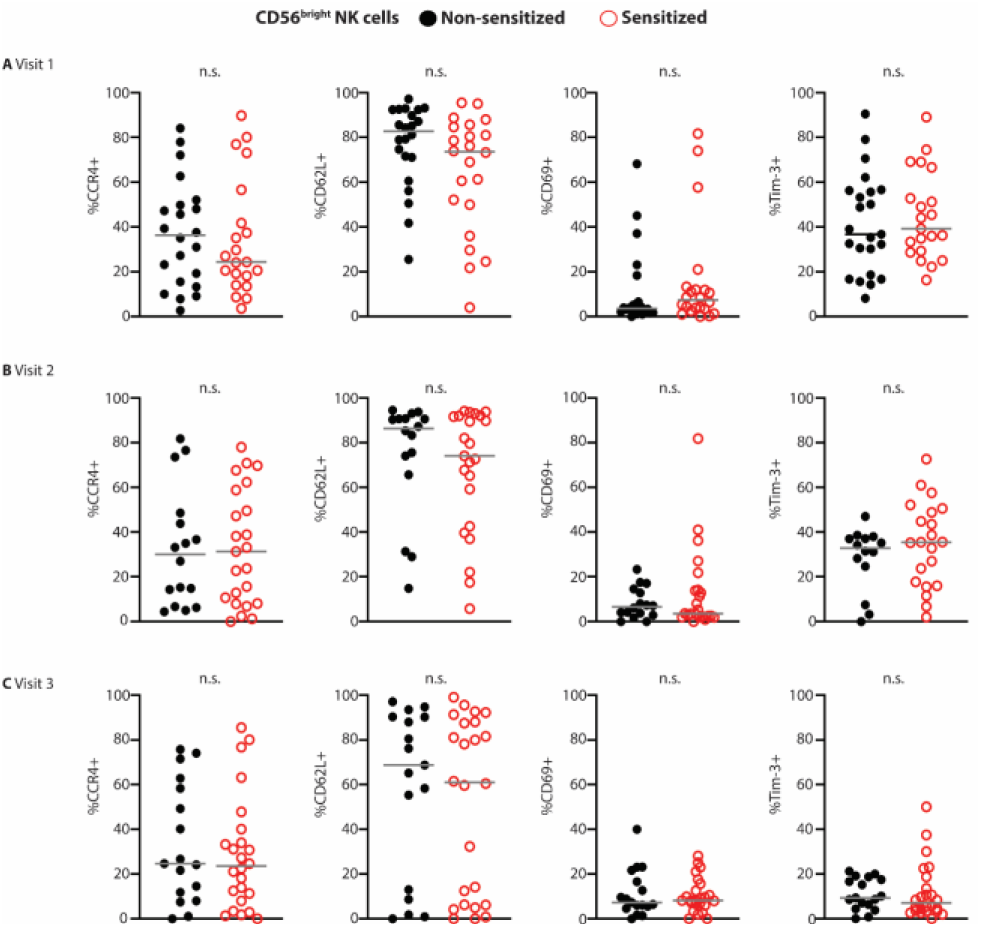
Allergen sensitization dependent changes in CD56^bright^ NK cell receptor expression across three annual visits. The expression of various surface markers on non-sensitized (closed black circles, V1, n=23; V2, n=16; V3, n=18) and sensitized (open red circles, V1, n=21; V2, n=23; V3, n=23) (**A-C**) Proportions of CD56^bright^ NK cells in each group of patients expressing CCR4, CD62L, CD69, and Tim-3 at V1-V3. Gray bars represent the medians in each group. Each point represents data from a single individual. Significant differences between group medians were determined by Mann-Whitney U test. No significant difference between medians was observed.

**Fig. S5.**
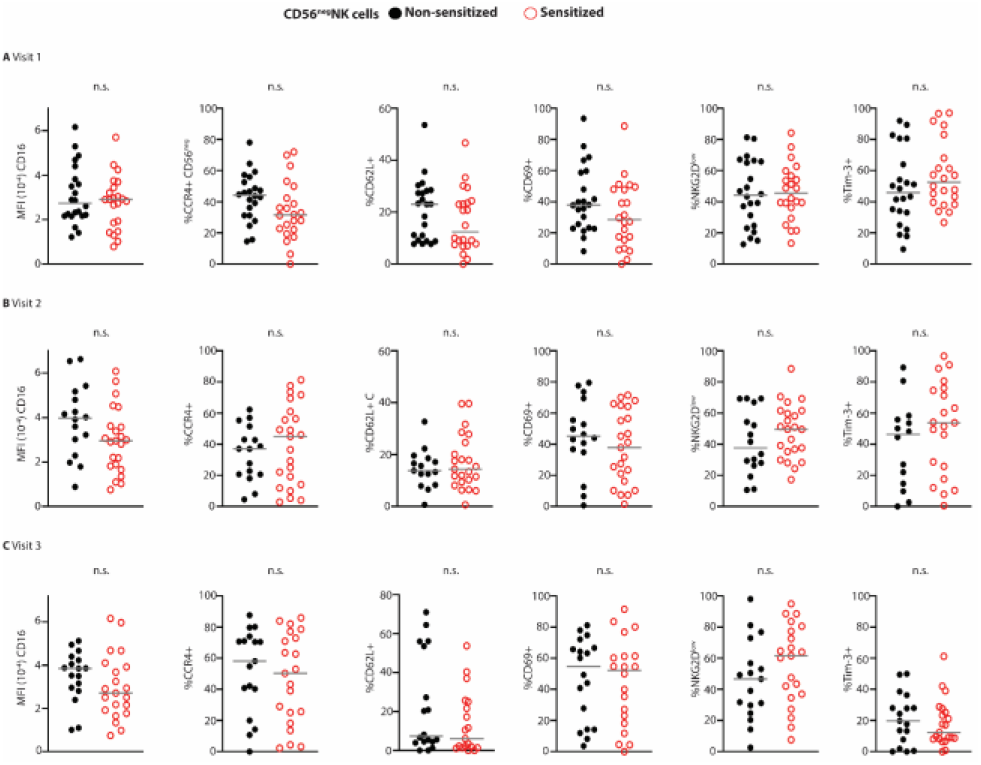
Allergen sensitization dependent changes in CD56^neg^ NK cell receptor expression across three annual visits. The expression of various surface markers on non-sensitized (closed black circles, V1, n=23; V2, n=16; V3, n=18) and sensitized (open red circles, V1, n=21; V2, n=23; V3, n=23) (**A-C**) Proportions of CD56^neg^ NK cells in each group of patients expressing CD16, CCR4, CD62L, CD69, and Tim-3 at V1-V3. Significant differences between group medians were determined by Mann-Whitney U test. Gray bars represent the medians in each group. Each point represents data from a single individual. No significant difference between medians was observed.

**Fig. S6.**
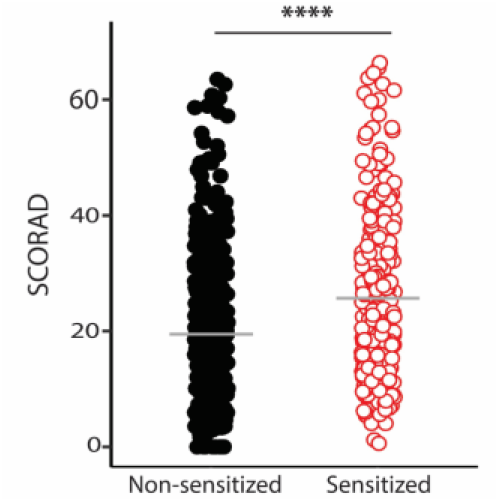
Children with allergen sensitization present increased AD severity. AD severity was assessed using SCORing for Atopic Dermatitis (SCORAD) for non-sensitized (closed black circles, n=387) and sensitized (open red circles, n=261) children. Gray bars represent the means in each group. Each point represents data from a single individual. Significant difference between group means was determined by two-sample t-test. **** p<0.001.

**Fig. S7.**
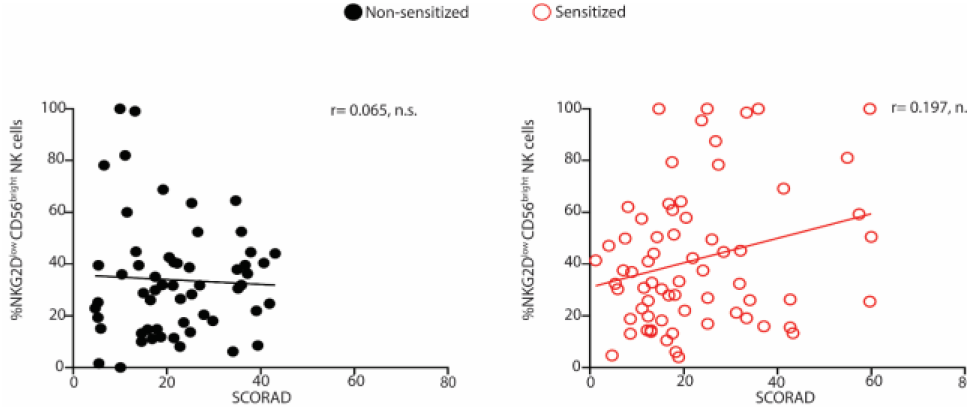
Association between CD56^bright^ NKG2D^low^ percentages to increased AD severity in sensitized children. Non-significant inverse relationships between SCORAD and proportion of NKG2D^low^ was observed in sensitized (open red circles, n=67) but not in non-sensitized (closed black circles, n=57) children (Combined from V1-V3) in CD56^bright^ NK cells. Spearman correlation test was used to assess association between SCORAD and NK cells. Each point represents data from a single individual.

**Figure S8.**
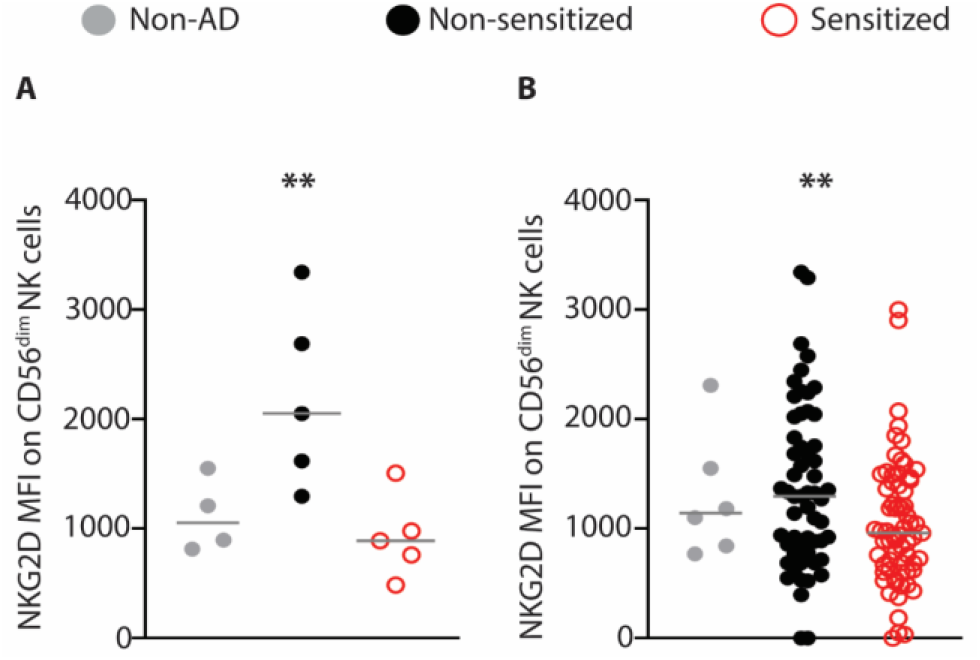
CD56dim NK cells from participants with reduced cytotoxicity show reduced NKG2D expression. **(A)** CD56dim NK cells derived from V1 and V2 for participants evaluated in Figure 4A+B without AD (closed gray circles, n=4) or MPAACH children with allergen sensitivity (open red circles, n=5) or non-sensitized (closed black circles, n=5) and were evaluated for MFI of NKG2D expression. **(B)** MFI of NKG2D on CD56dim NK cells was measured for children without AD (closed gray circles, n=6) or MPAACH children with allergen sensitivity (open red circles, n=67) or non-sensitized (closed black circles, n=57). Gray bars represent the medians in each group. Each point represents data from a single individual. Comparison of medians was done using Kruskal-Wallis test. ** p<0.05.

**Fig. S9.**
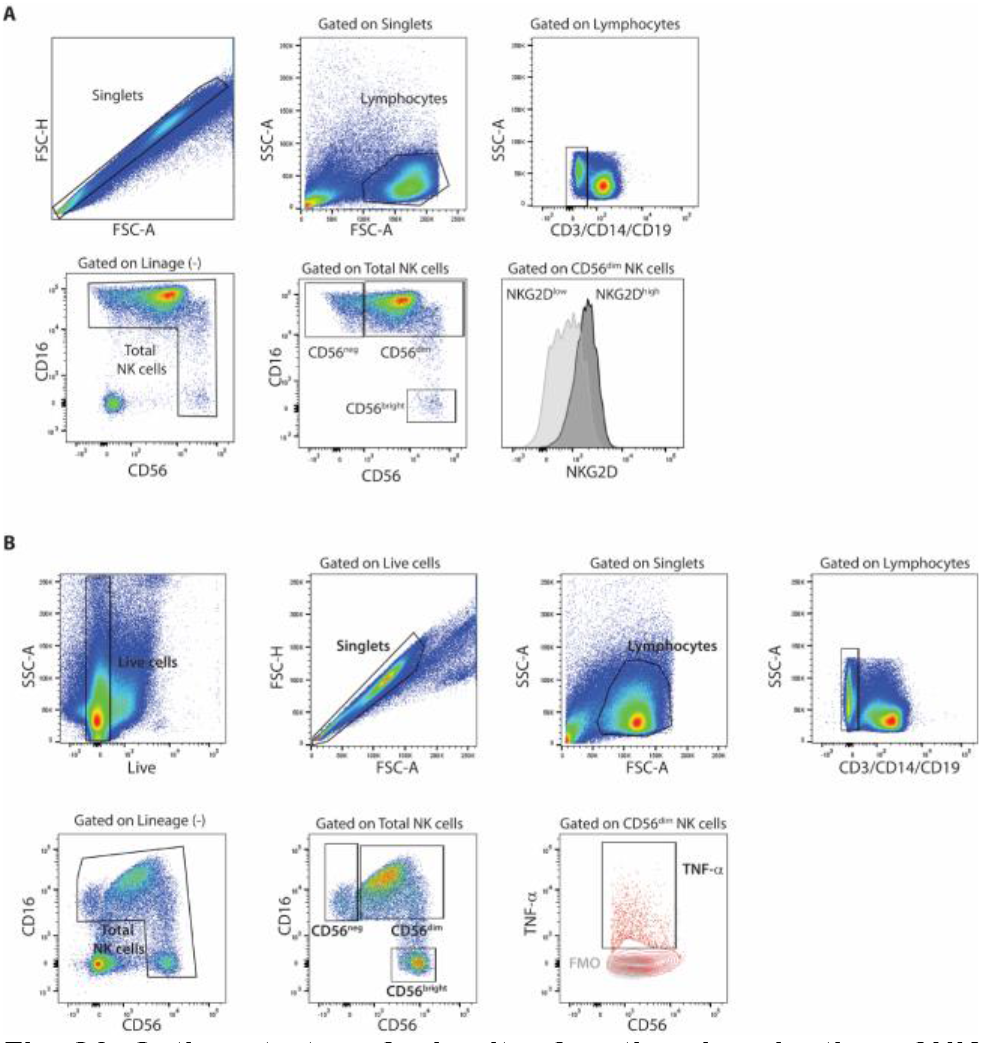
Gating strategy for in vitro functional evaluation of NK cells. Representative flow cytometry analysis of NK cells derived from **(A)** V3 analyzed in Figure 5 or **(B)** V1 and V2 in Figure 6A-D showing sequentially gating of live, singlet, live lymphocyte, and lineage negative (CD3neg CD19neg CD14neg) events prior to discrimination of CD16+CD56neg, CD16+CD56dim, or CD16neg CD56bright NK cells. This is followed by assessment of **(A)** NKG2D expression levels or **(B)** proportion of TNF-α+ among CD56dim NK cells (red) or FMO (gray).

**Fig. S10.**
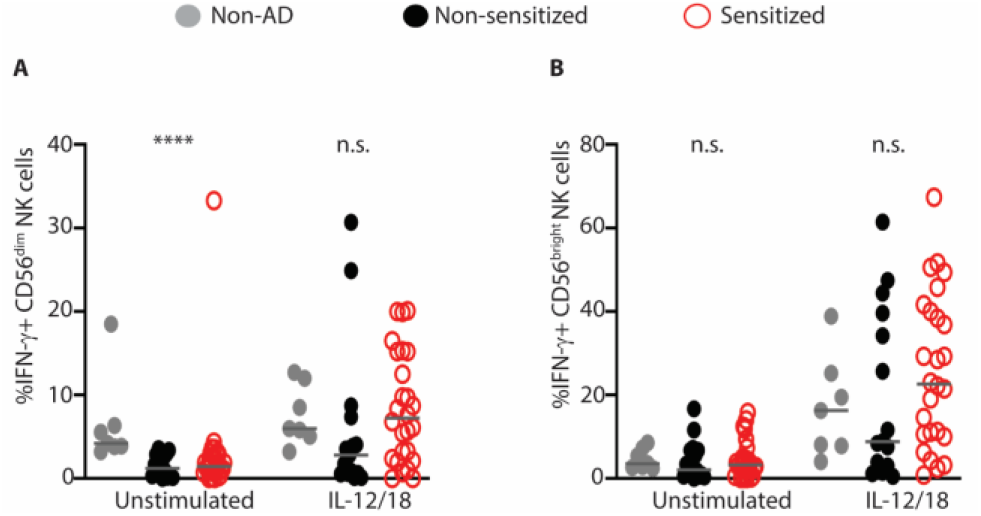
NK cells in allergen-sensitized patients show similar expression of IFN-γ to non-sensitized patients. Following culture of V1 and V2 PBMC in media (unstimulated) or IL-12 and IL-18, NK cells derived from non-AD (closed gray circles, n = 7), non-sensitized (closed black circles, n=15) and allergen sensitized (open red circles, n=26) children were evaluated for cytokine expression by flow cytometry. Expression of IFN-γ on (**A**) CD56^dim^ (**B**) CD56^bright^ NK cells. Gray bars represent the medians in each group. Each point represents data from a single individual. Comparison of medians was done using Kruskal-Wallis test. ****p<0.001.

**Fig. S11.**
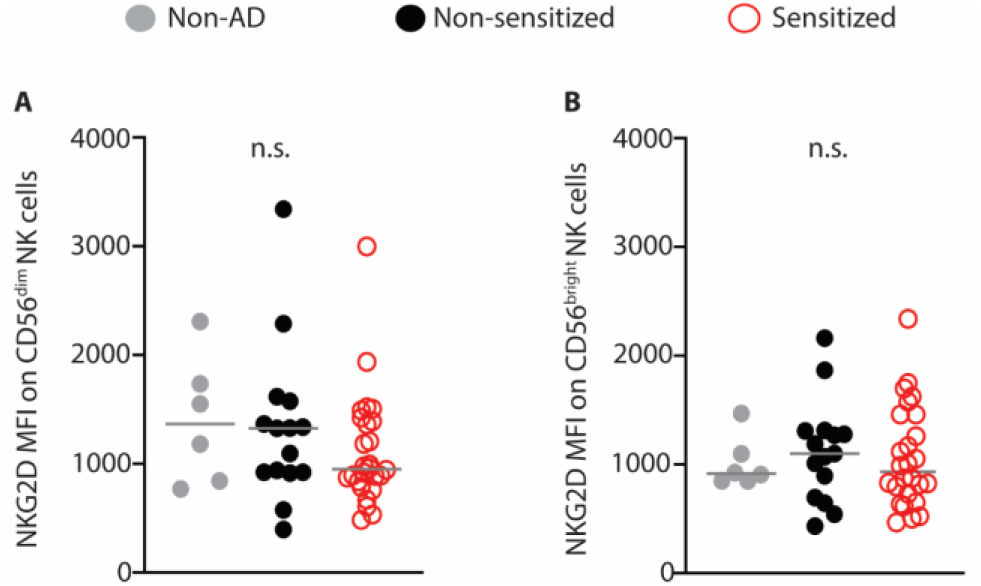
NKG2D expression on NK cells examined in cytokine expression assays. NK cells derived from Figure 6 participants (V1 + V2) including children without AD (closed gray circles, n=6) or MPAACH children without (closed black circles, n=15) or with (open red circles, n=26) allergen sensitivity, were evaluated for MFI of NKG2D expression on (**A**) CD56^dim^ and (**B**) CD56^bright^ NK cells. Gray bars represent the medians in each group. Each point represents data from a single individual. Comparison of medians was done using Kruskal-Wallis test. No significant difference between medians was observed.

**Fig. S12.**
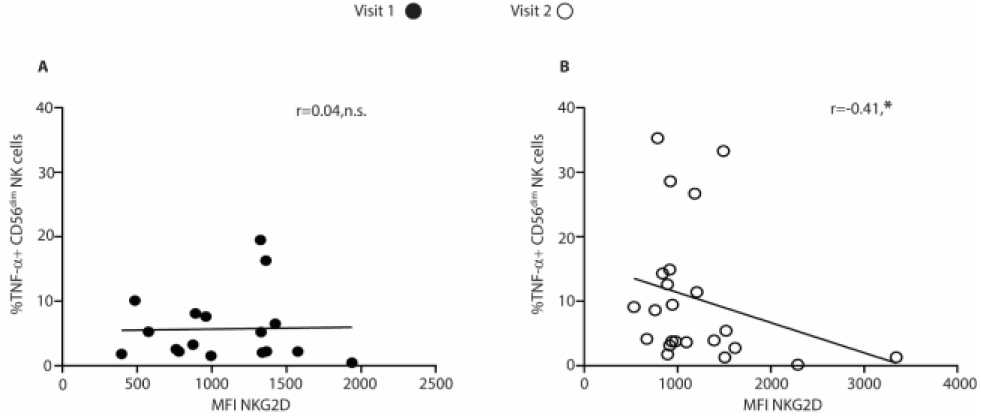
Negative correlation between NKG2D expression and proportion of TNF-α+ CD56^dim^ NK cells in children from Visit 2 but not Visit 1. An Inverse relationship between NKG2D mean fluorescence intensity and proportion of TNF-α+ CD56^dim^ NK cells was observed in V2 (open black circles, n=23) but not in V1 (closed black circles, n=18) children. Spearman correlation test was used to assess association between NKG2D expression to proportion of TNF-α+ CD56^dim^ NK cells. Each point represents data from a single individual. One V1 data point (MFI NKG2D = 3001; proportion of CD56^dim^ TNF-α+ = 28.6) was removed from the plot for better data visualization. * p<0.09.

**Fig. S13.**
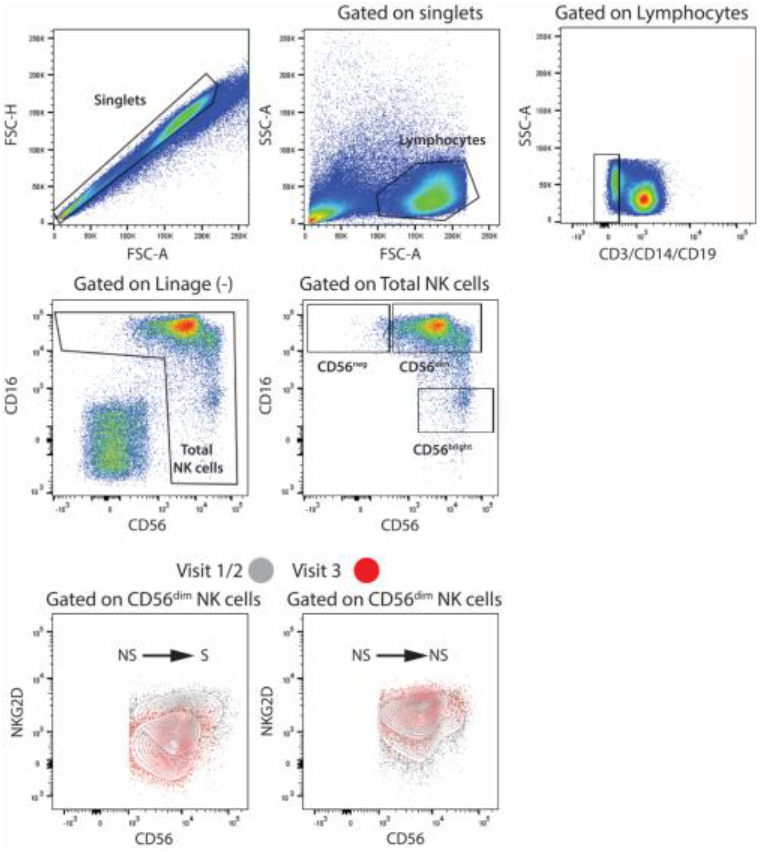
Gating strategy for longitudinal changes of NKG2D expression for NK cells in MPAACH cohort. NK cells derived from V1-V3 for participants evaluated in Figure 7A-D. Representative flow cytometry analysis with NK cells sequentially gated as, singlet, live lymphocyte, and lineage negative (CD3^neg^ CD19^neg^ CD14^neg^) events prior to discrimination of CD16^+^CD56^neg^, CD16^+^CD56^dim^, or CD16^neg^ CD56^brigh^t NK cells. NKG2D expression was evaluated on NK cells from V1/2 (gray) and V3 (red) within NS-S and NS-NS children.

**Fig. S14.**
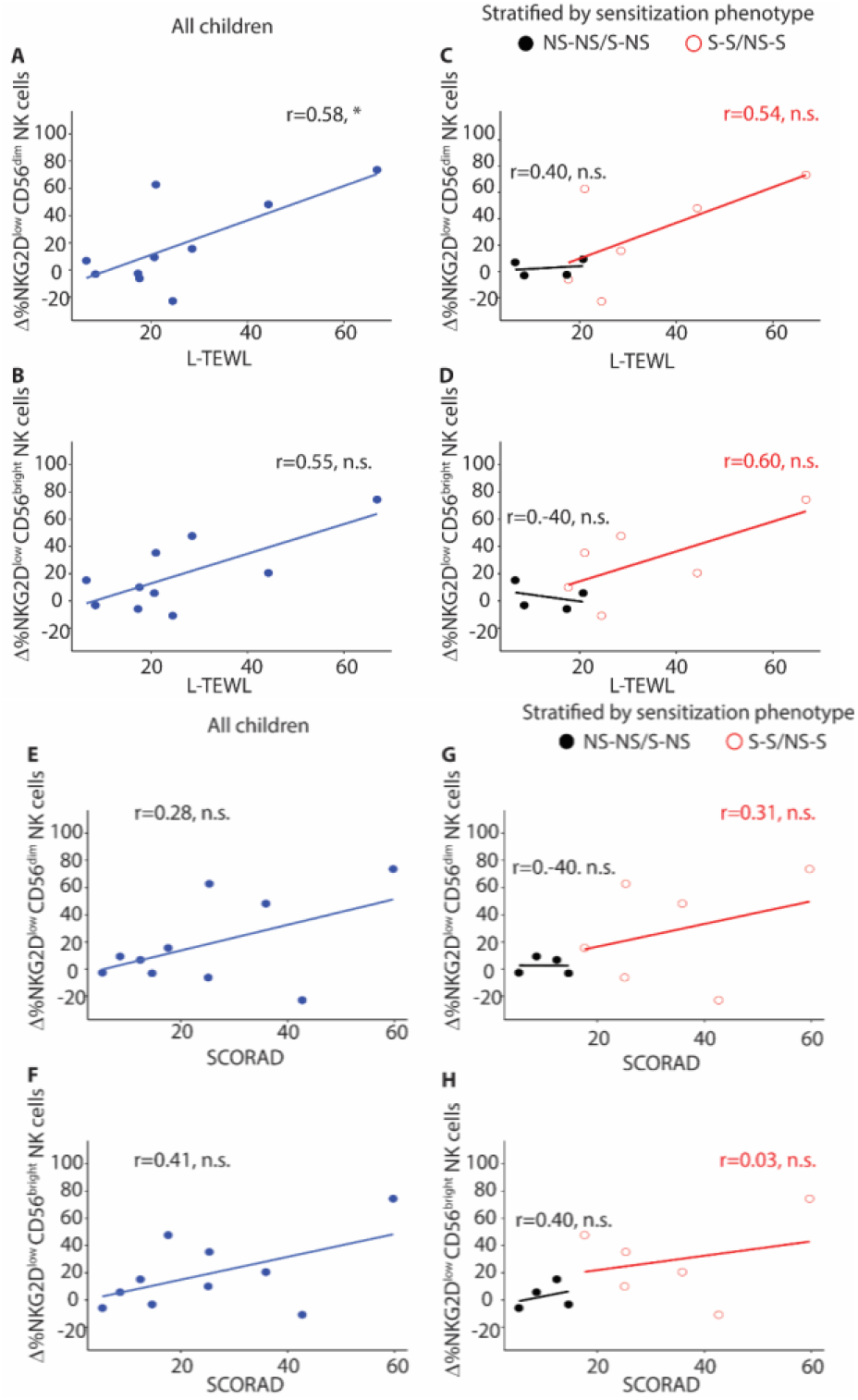
Association between changes in NKG2D expression over visits and skin barrier function and AD severity. **(A)** Association between L-TEWL and delta NKG2D^low^ CD56^dim^ and **(B)** delta NKG2D^low^ CD56^bright^ NK cells in children with longitudinal analysis of proportions of NK cells (all children, closed blue circles, n = 10), **(C-D)** stratified by sensitization phenotype (NS-NS/S-NS: closed black circles, n = 4; S-S/NS-S: open red circles, n = 6). **(E)** Association between SCORAD and delta NKG2D^low^ CD56^dim^ and **(F)** delta NKG2D^low^ CD56^bright^ NK cells, **(G-H)** stratified by sensitization phenotype. Association was assessed using Spearman correlation test. Changes (delta) in NKG2D^low^ CD56^dim^ and NKG2D^low^ CD56^dim^ NK cells were calculated as V3 – V1 or V2 NK cells expression. * p<0.09.

**Table S1.** Sample size and demographics in children by visit^1^.

|  | V1<br>n = 44 | V2<br>n = 39 | V3<br>n = 41 | P-value<sup>2</sup> |
| --- | --- | --- | --- | --- |
| **Demographic** |  |  |  |  |
| Age (y) | 1.8 (1.4-2.2) | 3.6 (3.4-3.7) | 4.8 (4.1-5.2) | < 0.001 |
| Male sex | 25 (57) | 23 (59) | 18 (44) | 0.34 |
| Black race | 27 (61) | 22 (56) | 22 (54) | 0.74 |
| Public insurance | 31 (70) | 21 (53) | 25 (61) | 0.49 |
| **AD Severity<sup>3</sup>** |  |  |  |  |
| SCORAD (score) | 18.6 (12.8-26.2) | 15.4 (9.6-26.4) | 20.0 (13.6-27.3) | 0.29 |
| Moderate/Severe AD (SCORAD ≥ 25) | 13 (29) | 11 (28) | 14 (34) | 0.88 |
| **Skin barrier function** |  |  |  |  |
| **Lesional skin** |  |  |  |  |
| TEWL (g/m<sup>2</sup>/h) | 11.2 (8.5-20.8) | 11.0 (7.5-16.5) | 16.6 (10.5-21.5) | 0.13 |
| Keratinocyte FLG expression | 1.46 (0.70-5.24) | 7.19 (2.89-11.32) | 3.37 (0.75-8.73) | 0.006 |
| **Non-lesional skin** |  |  |  |  |
| TEWL (g/m<sup>2</sup>/h) | 10.7 (7.7-13.9) | 9.6 (7.7-10.7) | 10.5 (8.4-14.9) | 0.11 |
| Keratinocyte FLG expression | 4.13 (1.23-11.75) | 6.25 (3.68-12.27) | 5.31 (1.30-10.60) | 0.29 |
| **Sensitization** |  |  |  |  |
| Total serum IgE | 32 (10-99) | 70 (24-143) | 87 (17-186) | 0.17 |
| Sensitization | 21 (48) | 23 (59) | 23 (56) | 0.70 |
| **Comorbid conditions** |  |  |  |  |
| Food allergy | 10 (23) | 10 (26) | 12 (29) | 0.79 |
| Wheezing in the past 12 mos | 14 (32) | 9 (23) | 19 (46) | < 0.001 |
| Allergies or allergic rhinitis | 13 (29) | 12 (31) | 23 (56) | 0.023 |

<sup>1</sup>Data are presented in median (IQR) or frequency (%). FLG expression was multiplied by 1000 for the ease of interpretation.
<sup>2</sup>Comparison across visits were done using Kruskal-Wallis or Fisher's exact tests.

**Table S2.**
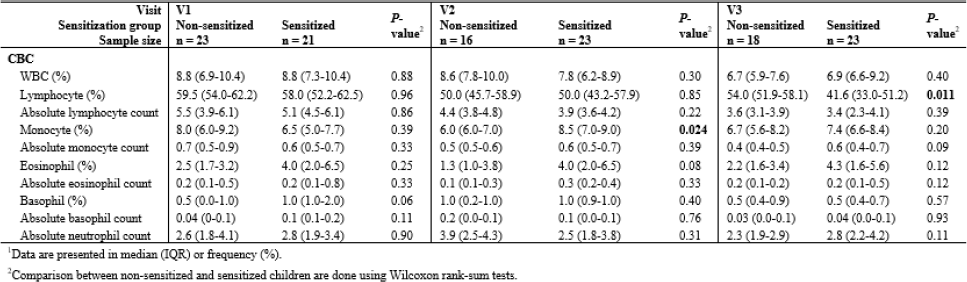
Complete blood count and cytokine level by sensitization^1^.

**Table S3.** Characteristics of the MPAACH cohort and the subset used in UMAP at visit I^1^.

|  | Full cohort<br>N = 651 | UMAP subset<br>n = 8 |
| --- | --- | --- |
| **Demographic** |  |  |
| Age (y) | 2.0 (1.2-2.4) | 2.4 (2.3-2.5) |
| Male sex | 345 (53) | 5 (62) |
| Black race | 444 (68) | 5 (62) |
| Public insurance | 457 (70) | 7 (87) |
| **AD Severity<sup>3</sup>** |  |  |
| SCORAD (score) | 19.5 (12.0-31.0) | 22.2 (17.8-38.27) |
| Moderate/Severe AD (SCORAD ≥ 25) | 202 (31) | 4 (50) |
| **Skin barrier function** |  |  |
| **Lesional skin** |  |  |
| TEWL (g/m<sup>2</sup>/h) | 12.4 (8.6-19.9) | 17.9 (15.8-29.1) |
| Keratinocyte *FLG* expression | 0.93 (0.25-2.40) | 2.93 (1.93-4.25) |
| **Non-lesional skin** |  |  |
| TEWL (g/m<sup>2</sup>/h) | 9.7 (7.4-13.4) | 10.2 (7.3-12.2) |
| Keratinocyte *FLG* expression | 1.51 (0.52-3.58) | 2.47 (1.37-3.81) |
| **Sensitization** |  |  |
| Total serum IgE | 30 (10-111) | 90 (35-141) |
| Sensitization | 261 (40) | 4 (50) |
| **Comorbid conditions** |  |  |
| Food allergy | 91 (14) | 0 |
| Wheezing in the past 12 mos | 241 (37) | 4 (50) |
| Allergies or allergic rhinitis | 102 (16) | 0 |

<sup>1</sup>Data are presented in median (IQR) or frequency (%). *FLG* expression was multiplied by 1000 for the ease of interpretation.

**Table S4.** Demographics by sensitization phenotype^1^.

|  | Non-/Transient Sensitization<br>n = 4 | Acquired/Persistent Sensitization<br>n = 6 | *p*<sup>2</sup> |
| --- | --- | --- | --- |
| **Demographic** |  |  |  |
| Age (y) | 2.1 (1.8-2.3) | 2.1 (1.6-2.5) | 0.91 |
| Male sex | 2 (50%) | 5 (83%) | 0.50 |
| Black race | 2 (50%) | 1 (17%) | 0.50 |
| Public insurance | 2 (50%) | 3 (50%) | 0.71 |
| **AD Severity** |  |  |  |
| SCORAD (score) | 10.5 (7.8-13.0) | 30.6 (25.2-41.0) | **0.009** |
| Moderate/Severe AD (SCORAD ≥ 25) | 0 (0%) | 5 (83%) | **0.048** |
| **Skin barrier function** |  |  |  |
| L-TEWL (g/m<sup>2</sup>/h) | 12.9 (8.0-18.2) | 26.5 (21.9-40.4) | **0.019** |
| NL-TEWL (g/m<sup>2</sup>/h) | 11.8 (9.0-14.7) | 13.9 (13.5-14.8) | 0.61 |

<sup>1</sup>Data are presented in median (IQR) or frequency (%).
<sup>2</sup>Comparisons between the two groups were done using Wilcoxon rank-sum and Fisher's exact tests.

## Notes

### Competing Interest Statement

The authors have declared no competing interest.

### Clinical Protocols

https://www.cincinnatichildrens.org/research/divisions/a/asthma/labs/hershey/current-projects/mpaach

### Funding Statement

Funding support provided by 5U19AI70235 (GKKH, JBM, LJM); American Heart Association (DEO); Asthma and Allergic Diseases Cooperative Research Centers (SA); R01AR073228 (SNW), 5T32GM063483 (SBD).

### Author Declarations

The Institutional Review Board of Cincinnati Children's Hospital Medical Center gave ethical approval for this work.

### Summary of Updates

The text, analysis, figures, and supplemental material have been updated and revised.

